# Understanding the Impact of Sociocultural Gender on Post-acute Sequelae of COVID-19: a Bayesian Approach

**DOI:** 10.1101/2021.06.30.21259757

**Authors:** Caroline E. Gebhard, Claudia Sütsch, Susan Bengs, Atanas Todorov, Manja Deforth, Karl Philipp Buehler, Alexander Meisel, Reto A. Schuepbach, Annelies S. Zinkernagel, Silvio D. Brugger, Claudio Acevedo, Dimitri Patriki, Benedikt Wiggli, Bianca Gysi, Jürg H. Beer, Andrée Friedl, Raphael Twerenbold, Gabriela M. Kuster, Hans Pargger, Sarah Tschudin-Sutter, Joerg C. Schefold, Thibaud Spinetti, Chiara Henze, Mina Pasqualini, Dominik F. Sager, Lilian Mayrhofer, Mirjam Grieder, Janna Tontsch, Fabian Franzeck, Pedro D. Wendel Garcia, Daniel A. Hofmaenner, Thomas Scheier, Jan Bartussek, Ahmed Haider, Muriel Grämer, Nidaa Mikail, Alexia Rossi, Núria Zellweger, Petra Opic, Angela Portmann, Roland von Känel, Aju P. Pazhenkottil, Michael Messerli, Ronny R. Buechel, Philipp A. Kaufmann, Valerie Treyer, Martin Siegemund, Ulrike Held, Vera Regitz-Zagrosek, Catherine Gebhard

## Abstract

**Background:** Women are overrepresented amongst individuals suffering from post-acute sequelae of SARS-CoV-2 infection (PASC). Biological (sex) as well as sociocultural (gender) differences between women and men might account for this imbalance, yet their impact on PASC is unknown.

**Methods and Findings:** By using Bayesian models comprising >200 co-variates, we assessed the impact of social context in addition to biological data on PASC in a multi-centre prospective cohort study of 2927 (46% women) individuals in Switzerland. Women more often reported at least one persistent symptom than men (43.5% vs 32.0% of men, p<0.001) six (IQR 5–9) months after SARS-CoV-2 infection. Adjusted models showed that women with personality traits stereotypically attributed to women were most often affected by PASC (OR 2.50[1.25-4.98], p<0.001), in particular when they were living alone (OR 1.84[1.25-2.74]), had an increased stress level (OR 1.06[1.03-1.09]), had undergone higher education (OR 1.30[1.08-1.54]), preferred pre-pandemic physical greeting over verbal greeting (OR 1.71[1.44-2.03]), and had experienced an increased number of symptoms during index infection (OR 1.27[1.22-1.33]).

**Conclusion:** Besides gender- and sex-sensitive biological parameters, sociocultural variables play an important role in producing sex differences in PASC. Our results indicate that predictor variables of PASC can be easily identified without extensive diagnostic testing and are targets of interventions aiming at stress coping and social support.

## Introduction

As the COVID-19 pandemic has evolved across the globe, male sex, cardiovascular and metabolic diseases and advanced age have been identified as predominant risk factors for a more severe disease course of COVID-19 and poor prognosis.(1) Accordingly, male patients with COVID-19 are reported to die at twice the rate of women when contracting the virus.(2) Biological (sex) differences in immune responses and expression levels of receptors responsible for viral entry and priming have initially been suggested to account for the higher COVID-19-related mortality rates seen in men.(3, 4) However, these findings have very recently been questioned or refuted.(5-7) In addition, factors beyond innate sex, such as institutionalized gender and culturally entrenched roles and norms, have been widely ignored in analyzing the causes of sex and gender disparities in COVID-19 outcomes, an omission that has been criticized by several institutions including the Canadian Institutes of Health Research and the European Parliament.(8, 9)

Increasing evidence suggests that severe acute respiratory syndrome coronavirus (SARS-CoV)-2 can cause a prolonged disease course beyond acute illness.(10-12) The clinical presentation of these post-acute sequelae of SARS-CoV-2 infection (PASC) includes a variety of fluctuating and unpredictable somatic symptoms persisting even beyond 12 months after initial infection, thereby posing a rising burden on healthcare systems.(13-18) In fact, predictions by the World Health Organization (WHO)(19) provide a grim perspective for the potential impact of the current SARS-CoV-2 Omicron variant that will infect up to 50% of Europeans in the first months of 2022. Therefore, increasing concerns are arising that PASC could soon become the leading chronic disease in Western countries.

Although morbidity from acute COVID-19 infection is substantially lower in women than men, women are overrepresented amongst patients suffering from PASC.(20-24) Accordingly, factors increasing the risk of severe acute COVID-19 illness, such as advanced age or male sex, do not also increase the risk of PASC.(25) The causes for the differential sex- and gender distribution in acute versus chronic COVID-19 illness remain enigmatic. Given the currently unexplained gender disparity in PASC, the paucity of sex-disaggregated data, and the complete lack of reports considering gendered psychosocial variables beyond clinical-biological parameters in PASC, we sought to assess the impact of social context, gender, and behaviors in addition to biological data on PASC in a large multi-center cohort in Switzerland comprising both hospitalized patients and outpatients with confirmed SARS-CoV-2 infection. In light of the many unknown variables that might contribute to the risk of PASC as well as the challenge to include gender, a complex construct, in the analysis, Bayesian modelling was chosen as novel statistical approach to perform a gender-sensitive analysis.

## Methods

### Study design and procedures

This study is based on data from patients of the Swiss COGEN cohort study, a prospective, observational cohort of polymerase chain reaction-confirmed SARS-CoV-2 infected individuals diagnosed between February and December 2020 at one of four Swiss study sites. Eligible patients were adults aged ≥18 years at follow-up who survived acute COVID-19 infection, residing in Switzerland during primary SARS-CoV-2 infection, and fluent in German, English, French, or Italian and able to provide informed consent. After a minimum follow-up time of 60 days, each participant was contacted by telephone and asked to complete a questionnaire either by phone, email, or written form. Out of 6028 patients, 3009 patients or their legal representatives completed the questionnaire after giving informed consent (**Supplemental Figure 1)**. Additional details on study design and procedures are provided in **Supplemental Methods**.

### Ethics Approval

The study complies with the Declaration of Helsinki and its later amendments, and the research protocol was approved by the responsible ethics committee of the Canton of Basel (EKNZ, ethics approval #2020-01311). Informed consent was obtained from all patients or their legally authorized representative, as appropriate.

### Outcome measures

The primary outcome measure of our analysis was defined as the persistence of at least one COVID-19 related somatic symptom beyond 60 days after index infection. This definition was used by considering the varying classifications of PASC issued by the National Institute for Health and Care Excellence (NICE), the National Institutes of Health (NIH), and the US Centers for Disease Control and Prevention (CDC), which are based on constantly evolving epidemiological understanding of post- and long-COVID-19. The secondary outcome measure consisted of a group of *specific* persisting somatic symptoms of COVID-19. The latter comprised a cluster of somatic symptoms including dyspnea, reduced exercise performance, and changes in smell and taste, which have previously been shown to be distinctive features of COVID-19 in both women and men.(26-30) Further details are given in **Supplemental Methods**.

### Gender score and statistical analyses

The assessment of gender requires a complex approach which is described in detail in **Supplemental Methods** and **Supplemental Figure 2**. Statistical analysis was performed using R version 4.1.0 including the packages rstanarm and MCMCvis.(31) Simple group comparisons were performed using t-test, non-parametric rank sum test, or chi-square test as appropriate. Standard logistic regression was performed to assess the predictive strength of biological (binary) sex and a summary gender score (see description in **Supplemental Methods**) for the persistence of any symptom (primary study outcome) or a cluster of specific symptoms (secondary study outcome) after a minimum of 60 days following acute SARS-CoV-2 infection. In each case, the model was corrected for age, recruitment site, overall QoL of each participant, and the prevalence of COVID-19 in Switzerland at the time of diagnosis. Models were compared based on the Nagelkerke pseudo-R^2^. Bayesian models were used to estimate the probability of the primary and secondary study outcomes. A detailed description of the Bayesian models as well as an illustration of the statistical approach is provided in **Supplemental Methods** and **Supplemental Figure 3**. Notably, we consciously decided not to perform causal modelling to avoid bias in our causal framework and focus instead on identifying the best predictive model by using all available data. Although we did not perform a specific confounder analysis per variable of interest, our prediction models were de-confounded to the highest possible degree by only considering coefficients derived by multivariate models.

## Results

### Biological sex and its association with PASC

During a median follow-up time of 6 ([IQR] 5-9) months (p=NS for males vs females) 1091 (37.3%) individuals reported at least one somatic symptom (primary outcome measure) that persisted following primary infection. Females more often reported at least one persistent somatic symptom as compared to males (43.5% vs 32.0%, p<0.001, **Figure 1A and Supplemental Figure 4**). The higher prevalence of PASC in females was observed in both outpatients (40.5% in females vs 25.4% in males, p<0.001) and hospitalized patients (63.1% in females vs 55.2% in males, p<0.001). The prevalence of PASC was higher in hospitalized patients than in outpatients (57.9% vs 32.7%, p<0.001, **Figure 1A** and **1B**).

**Figure 1:**
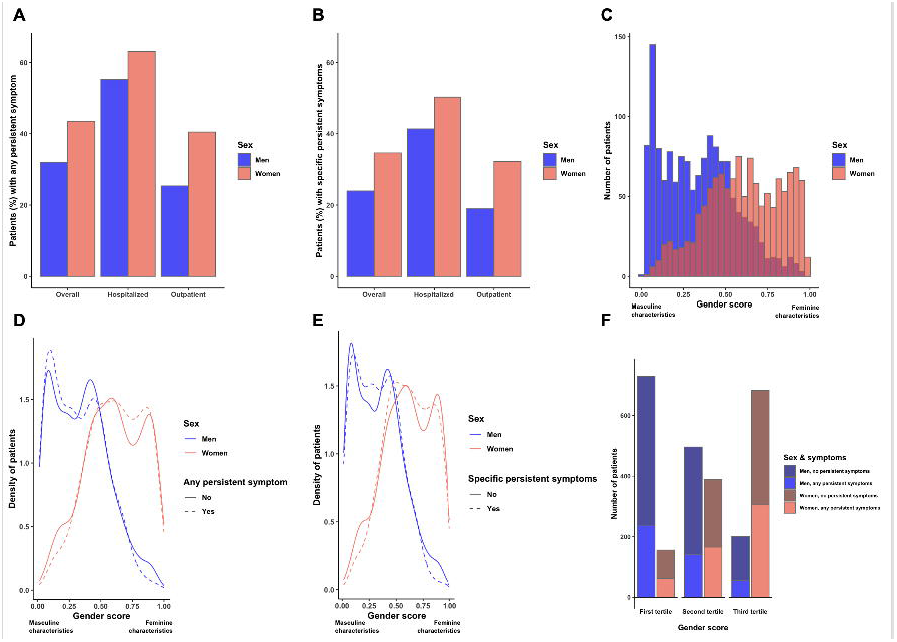
**(A)** Percentage of patients reporting **any somatic symptom** in overall study population (left), hospitalized patients (middle), and outpatients (right) stratified by sex as a binary variable. **(B)** Percentage of patients reporting **at least one specific somatic** symptom in overall study population (left), hospitalized patients (middle), and outpatients (right) stratified by sex as a binary variable. **(C)** Distribution of the gender score stratified by sex as a binary variable (male/female). Gender variables included in the score covered gender roles, gender-relations, and institutionalized gender. The selected variables were measured using self-report-questionnaires and the score was constructed via logistic regression. The resulting gender score ranged from 0.00-1.00, with higher values reflecting more traditionally feminine characteristics and lower values more traditionally masculine characteristics. **(D)** Association of both, the gender score and sex as a binary variable with the primary endpoint of the study (**persisting symptoms** following SARS-CoV-2 infection). **(E)** Association of both, the gender score and sex as a binary variable with the secondary endpoint of the study (**persisting specific somatic symptoms** following SARS-CoV-2 infection). **(F)** Number of patients reporting **no** and **any persistent symptom** stratified by gender score (tertiles) and sex as a binary variable. Data are presented as percentage of all women (n=583) and men (n=508) reporting any persistent symptom **(A** and **B)**, absolute numbers **(C)**, or median/interquartile and total range/outliers **(F)**.

### Gender score and its association with PASC

The calculation of the gender score is described in detail in **Supplemental Methods** and **Supplemental Figure 2. Figure 1C** shows the distribution of the gender score for men and women separately. As expected, men on average scored lower while women scored higher. Women with characteristics traditionally ascribed to women (3^rd^ tertile of the gender score) were more likely to experience PASC than women with a lower gender score (1^st^ tertile of the gender score, **Figure 1D and 1F**). Similar, men with characteristics traditionally ascribed to men (1^st^ tertile of the gender score) were more likely to report persistent symptoms than men with a higher gender score (3^rd^ tertile, **Figure 1D** and **1F**).

### Biological sex and its association with *specific somatic symptoms* of PASC

When only a cluster of specific somatic symptoms was considered, females reported more often than males the persistence of these symptoms (34.6% vs 23.9%, p<0.001, **Figure 1B**).

### Gender score and its association with *specific somatic symptoms* of PASC

The associations between gender score and PASC did not alter when only the persistence of *specific somatic* symptoms was considered (**Figure 1E**).

### Comparison of gender score versus sex in the prediction of PASC

Prognostic models for PASC adjusted for age, recruitment site, overall QoL of each participant, and the prevalence of COVID-19 in Switzerland at the time of diagnosis and including either biological sex or gender score showed that each of these variables could significantly predict PASC (p<0.0001, **Table 1, upper panel** for any symptom and **lower panel** for *specific somatic* symptoms).

### Bayesian models: overall population

#### Biological and sociocultural determinants of PASC

Bayesian models were used to estimate the probability of PASC in the overall population by including all 249 variables without preselection as well as interaction terms consisting of biological sex*variable and gender score*variable in the model. Individuals who reported at least one persistent COVID-19 related somatic symptom (primary outcome) were more likely to be widowed and women (odds ratio [OR] and 90% credible interval [CI] 3.13[1.82-5.45]), obese (OR 1.67[1.39-2.00]), had more often dyspnea at presentation for primary infection (OR 1.57[1.34-1.84]), suffered more often from more than one unspecific (OR 1.27[1.22-1.33]) or specific somatic symptom (OR 1.07[1.00-1.15] during primary infection, had a higher education level (OR 1.29[1.08-1.54]) or reported more often an increased domestic stress-level (OR 1.06[1.03-1.09], **Figure 2A**). Characteristics typically ascribed to women in combination with gastrointestinal (GI) symptoms (OR 1.70[1.28-2.23]) or cough (OR 1.38[1.13-1.69]) at presentation for primary infection were also associated with an increased risk of persistent symptoms (**Figure 2A)**.

**Figure 2:**
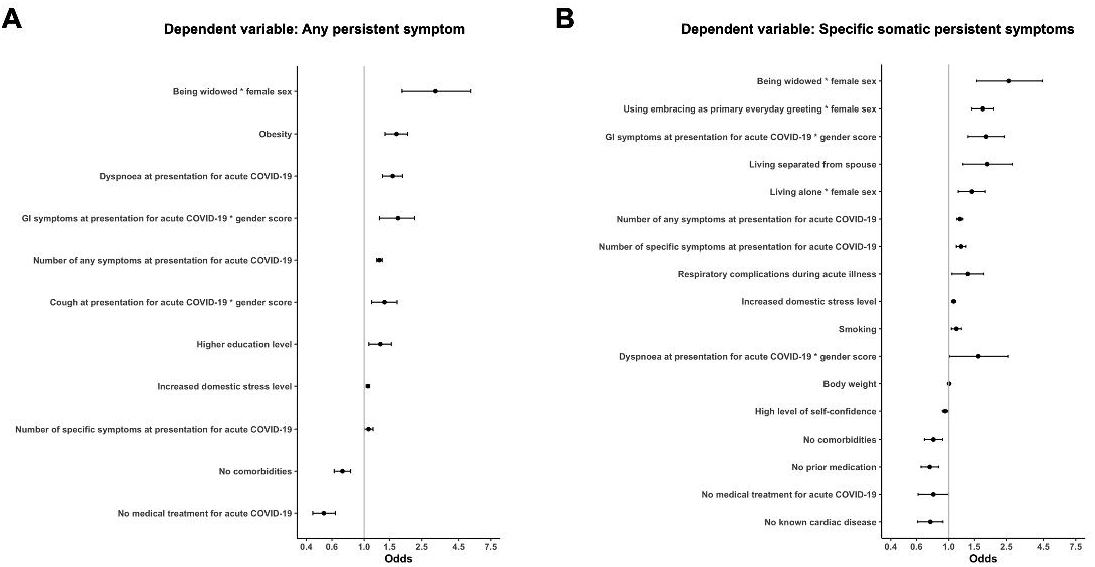
Forest Plot depicting log odds ratios (median and 90% credible intervals) of risk/protective factors associated with **(A)** any persistent symptom and **(B)** specific somatic persistent symptoms following SARS-CoV-2 infection in the total study population. GI, gastrointestinal.

#### Biological and sociocultural determinants of *specific somatic symptoms* of PASC

Similarly, individuals who reported a cluster of specific somatic symptoms (secondary outcome) were more likely to be women and widowed (OR 2.60[1.55-4.43]) or living alone (OR 1.44[1.16-1.78]), to live separated from spouse (OR 1.84[1.25-2.74]), or to have experienced more than one specific somatic (OR 1.21[1.13-1.31]) or unspecific symptom (OR 1.19[1.14-1.25]) at presentation for primary infection (**Figure 2B**). In addition, women who had used hugging as primary pre-pandemic greeting (OR 1.71[1.44-2.03]) were more likely to experience PASC. Characteristics typically ascribed to women in combination with GI symptoms at presentation for primary infection were amongst the strongest predictors of persistent specific symptoms (OR 1.80[1.36-2.41]). Weaker predictors of persistent specific symptoms were an increased domestic stress level (OR 1.08[1.05-1.11]), respiratory complications during primary infection (OR 1.35[1.05-1.73]), smoking (OR 1.13[1.04-1.22]), or characteristics typically ascribed to women in combination with dyspnea at presentation for acute COVID-19 (OR 1.60[1.01-2.56], **Figure 2B**). **Supplemental Figure 5** visualizes some of the interaction terms showing a strong association with PASC. A detailed description of these interactions with sex and gender score is provided in **Supplemental Results**.

### Bayesian models: hospitalized patients

#### Biological and sociocultural determinants of PASC

Bayesian models were used to estimate the probability of PASC in hospitalized patients by including 147 variables (all variables selected by the model for the overall population plus all available clinical and laboratory parameters) as well as interaction terms including sex and gender score in the model. Amongst hospitalized patients (n=532, 66.4% men and 33.6% women) being widowed (OR 4.51[2.08-9.58]) or presenting with cough at index infection (OR 1.73[1.13-2.71]) were the strongest predictors of PASC, followed by a higher gender score in combination with lower oxygen saturation at the first day of hospitalization (OR 1.60[1.30-1.96]), having more than one symptom at presentation for acute COVID-19 (OR 1.44[1.31-1.59]), and an increased domestic stress level (OR 1.19[1.04-1.35], **Figure 3**). Interaction terms selected by the model are depicted and described in **Supplemental Figure 6** and **Supplemental Results**.

**Figure 3:**
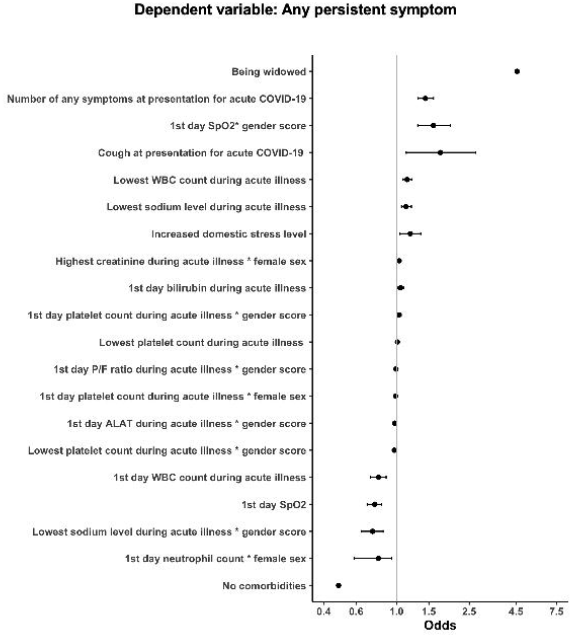
Forest Plot depicting log odds ratios (median and 90% credible intervals) of risk/protective factors associated with **any persistent** symptom following SARS-CoV-2 infection **in hospitalized patients**. SpO_2_, oxygen saturation; WBC, white blood cell; P/F ratio, PaO2/FiO2 ratio; ALAT, alanine aminotransferase.

### Selected patient characteristics stratified by sex and the presence/absence of PASC

The final study cohort comprised 2927 individuals of (1340[45.8%] females and 1587[54.2%] males, **Table 2**) and was stratified by biological sex and the presence or absence of PASC.

#### Biological variables

The median age (interquartile range [IQR]) of the study sample was 39[28-54] years. The average number of comorbidities was higher in men with PASC as compared to women with PASC ([mean±SD] 1.4±1.7 vs 1.1±1.5, p<0.001), while this difference was not observed in individuals without PASC (0.7±1.1 vs 0.6±1.0, p=0.106, **Table 2, Supplemental Figure 7A**). The average number of reported symptoms during primary infection was higher in females as compared to males, and higher in individuals with PASC as compared to those without PASC (absence of PASC: 3.9±2.1 in males vs 4.6±2.2 in females, p<0.001; with PASC: 5.3±2.3 in males and 6.0±2.3 in females, p<0.001, **Supplemental Table 1**).

#### Sociocultural variables

Women with PASC had more often obtained a university or technical college degree than men in this group, however, these differences were not statistically significant (42.9% vs 37.6% in men, p=0.15, **Supplemental Table 2**). Women were more often single parents than men, with the highest percentage of single parent women being observed in the PASC group (12.2% women vs 6.7% men, p<0.001). Also, women with PASC were more often divorced/separated (9.3% vs 7.3% in men, p=0.03), more often single (20.2% vs 15.6%, p=0.03) were more often widowed (4.8% vs 3.0%, p=0.03, **Supplemental Figure 7B** and **7C**), and less often married/living in a partnership than men (65.5% vs 73.6%, p=0.029 for men vs women, **Supplemental Table 2**).

A detailed description of the study cohort including clinical, inflammatory, and routine laboratory parameters during index infection, PASC characteristics as well as sex-specific and socioeconomic variables, is provided in **Table 2, Supplemental Tables 1-7** and **Supplemental Results**.

## Discussion

Our study is the first to report that, besides gender- and sex-sensitive biological parameters, sociocultural variables play a major role in producing sex differences in PASC. Women with typical feminine personality traits were most affected by PASC, in particular when they were living alone, had an increased stress level, higher education, had used hugging as primary pre-pandemic greeting, and had presented with a higher number of symptoms, specifically GI symptoms, during primary COVID-19. Our study adds to increasing evidence indicating that COVID-19 sex disparities cannot solely be explained by sex-specific biological mechanisms and are also explained by gendered or sex-specific patterns in health behaviors, contextual factors, and pre-existing health conditions.(7) In line with this notion, sex hormone status, hormone replacement, or hormone deprivation therapies were not associated with PASC in either sex in our study, although it has recently been proposed that the symptoms of PASC may overlap with those of perimenopause.(32) Similarly, psychosocial and behavioral factors and their interaction with sex/gender remained amongst the strongest predictors of PASC, even when only hospitalized patients were analyzed who, amongst all study groups, were best characterized regarding biological (clinical and laboratory) variables. Consistent with our conclusion, substantial variation in the magnitude and direction of COVID-19 sex disparities across geographical localities, amongst racial and ethnic groups, and over time exists, all of which are better explained by contextual factors than biological variables.(7, 33, 34)

The risk predictors for PASC selected by our models confirm previous reports indicating that psychosocial stress has substantially increased in women during the pandemic. Indeed, women have been disproportionately affected by imposed quarantine and lockdown measures given that typical feminine roles such as parenting, home-schooling, and other caring duties are still predominantly assumed by women.(35) Isolation at home measures along with financial and security concerns can put an additional strain on families, which in some situations can lead to high stress levels in women. Accordingly, women with PASC in our study reported the highest domestic stress level amongst all study groups, and the latter was a significant predictor of PASC in our models. On the other hand, women’s economic participation and educational attainment has continuously increased during the last two decades, which is reflected by the high number of women with a university or college degree in our population. Having less time for education, work, and career advancement, can lead not only to significant psychosocial stress, but also to increased professional and social inequality as it has recently been shown in women scientists.(36) Hence, the fact that women with PASC had more often obtained a university degree than men, a sex difference that was inversed in individuals free of PASC, might not only be reflective of a widening gender gap during the pandemic, but also of an exacerbation of role conflicts and subsequent detrimental effects on women’s health. Our observation that a higher education level was associated with an increased risk of PASC mirrors not only the vulnerable socioeconomic positions of women during the pandemic but also highlights that their ability to return to work might further be impeded by the chronicity of symptoms of PASC.

We observed a gendered relationship between the number and type (GI symptoms, cough, or dyspnea) of acute-phase symptoms, recorded at presentation for primary infection, and the risk of PASC months later. In addition, the number of symptoms during both acute illness and PASC showed a significant and positive correlation with the gender score in our study, indicating that individuals with more feminine characteristics reported in general a higher number of symptoms. While a higher degree of self-reflection and introspection in women might, at least in part, account for their higher number of reported symptoms,(37) the underlying mechanisms accounting for these association remain unknown. Nevertheless, in line with a recent report from Perlis et al.,(38) we observed a significant and positive correlation between the number of acute symptoms and the presence of anxiety, stress, or depression at follow-up in both women and men, thereby emphasizing the importance to consider potential neuropsychiatric sequelae of COVID-19.

It is also notable that the presence of GI symptoms during primary infection was amongst the strongest predictors of PASC. GI symptoms were more often observed in women than in men and showed a positive interaction with feminine gender in the prediction of PASC. GI symptoms are highly prevalent in COVID-19 ranging from 17.6% to 53%, and the high abundance of the virus entry receptor angiotensin-converting enzyme-2 (ACE-2) in the GI tract has been suggested to account for this finding.(39-41) Previous work also suggests a longer duration of illness, but no increased mortality in patients with GI symptoms, which aligns with our findings.(42) Delayed diagnosis and treatment of acute COVID-19, inflammation-mediated tissue damage in the gut, alterations of gut microbiota by the virus, an involvement of the brain-gut axis resulting in an increased risk of somatoform disorders have all been suggested to account for acute and long-term sequelae of COVID-19 in the GI tract.(42-45) Although sex-disaggregated data are lacking, sex differences in intestinal ACE-2 expression(46) or a female predisposition for certain GI disorders such as the irritable bowel syndrome, which is 2-3 times more common in working women,(47, 48) might account for the gendered impact of GI symptoms on PASC. Clinical trials assessing different treatment options of long-term sequalae of COVID-19 infection in the GI tract are currently ongoing and may shed light on these associations.

Consistent with current epidemiological evidence, women more often than men led single-parent families or lived alone in our study. The latter was a significant predictor of PASC in women across different subgroups and statistical models in our study. Isolation and the reduction of social relations together with economic tension has affected the general population but may even impose a larger burden on female COVID-19 survivors. In accordance with this assumption, women, individuals with advanced education degrees and/or chronic conditions as well as individuals living alone were more likely to report a feeling of social isolation, loneliness, or depression during the pandemic.(49-51) Similarly, COVID-19 mitigation and control strategies, like lockdowns and social distancing, have led to increases in psychological stress, depression, and anxiety, and reductions in general well-being, particularly in young adults and women.(52) Very frequent in-person connections were generally associated with lower depression and loneliness.(51) However, women are more likely to perceive COVID-19 as a very serious health problem, to agree with restraining public policy measures and to change behavior in response to new risk.(53) Consequently, behavioral changes in person-to-person contact might affect women more than men. Indeed, having used embracing as primary pre-pandemic everyday greeting was a significant predictor for PASC in our study. Given that women in general use embracing as greeting more often than men,(54) it is conceivable that limited person-to-person contact due to social distancing measures imposes additional stress and burden on women, thereby exacerbating symptoms of PASC.

The strength of our analysis consists of its near-complete, geographically defined cohort with availability of more than 200 clinical, laboratory, socioeconomic and psychosocial variables, the capture of a wide spectrum of post-COVID-19 symptoms, the availability of SARS-Co-V2 swab test in all study participants, the multicentre design permitting to include both outpatients with mild disease as well as hospitalized patients. Given the many unknown factors that might contribute to the risk of PASC, we have chosen Bayesian modelling as statistical approach which allowed us to include many variables in the model thereby avoiding the risk of a potential variable preselection bias. In addition, our models were corrected for study site, overall QoL, and prevalence of COVID-19 infection in Switzerland during the time of the study. Data characterizing primary infection were collected during ambulatory visits or hospitalization thereby minimizing recall bias. However, our study also has several limitations related to its cross-sectional and observational design. First, although the variables in our study covered many aspects of sex- and gender-specific demographic, behavioral and contextual characteristics, residual confounding due to unmeasured parameters in our dataset is possible. Second, self-selection or other biases may have occurred if individuals who are more concerned with their health were more likely to participate. Third, our study was conducted in Switzerland, a high-income country with low gender-inequality index. Given that gender-related characteristics are culturally sensitive, our observations may not be extrapolated to other societies and geographical regions. Fourth, participants who did not complete follow-up were less likely to have comorbidities than those with follow-up data. Given that the absence of comorbidities was more common in women and was associated with a reduced risk of PASC, the inclusion of patients with missing follow-up would have likely decreased the prevalence of PASC in the overall population. Finally, to date there is no academic consensus on how to define the construct “gender”. Consequently, there is no gold standard for a measure of gender and as such, our gender-related score is internally derived without external validation. Also, the gender score was developed and validated in Canada and Germany, thus, the behaviours ascribed to women and men reflect a Western understanding based on data from upper income countries.

Taken together, while biological variables may have a role in explaining sex disparities in COVID-19, our study also suggests a major impact of societally constructed characteristics of men and women in producing sex and gender differences in PASC. Currently, the CDC suggest a symptom-specific approach to treat PASC, however, our data indicate that post-COVID-19 sequalae and their predictors differ between men and women suggesting that a tailored and sex- and gender-sensitive approach of healthcare services may be required to support their needs. Indeed, many predictor variables of PASC identified in the present study are modifiable and targets of interventions aiming at stress coping and social support. Also, the reported PASC risk factors can be easily identified at an early stage of disease by taking a thorough patient history without additional blood sampling or extensive diagnostic testing. The latter allows to enable preventive measures already during acute-phase infection. Further research will be needed to determine if interventions targeted at these factors could improve outcomes.

## Data Availability

Based on the Business Administration System for Ethics Committees (BASEC) ethics approval, the non-anonymized raw data cannot be shared publicly. However, anonymised data that underlie the results reported in this article will become available to interested parties for non-commercial reasons, after the publication upon reasonable requests made to the corresponding author. Data requestors will need to sign a data access agreement.

## Acknowledgement

We would like to thank the staff of the Department of Nuclear Medicine of the University Hospital Zurich and the Intensive Care Units of the University Hospital Basel and the University Hospital Zurich for their excellent work and dedication to this study. The authors thank Christian Schindler, PhD, biostatistician at the Swiss Tropical and Public Health Institute Basel, Switzerland, for providing expert advice to the statistical analysis and for critically reviewing the manuscript. And last, we thank all the study patients for their valuable time and commitment to the Swiss COGEN Cohort.

## Declarations

### Transparency statement

The manuscript’s guarantor (CG) affirms that the manuscript is an honest, accurate, and transparent account of the study being reported; that no important aspects of the study have been omitted; and that any discrepancies from the study as originally planned have been explained.

### Contributor information

CG, CEG, and VRZ conceptualized and designed the Swiss COGEN study. CEG, CS, SB, KPB, NH coordinated the study. CEG, PO, SB, AM, NZ prepared the study data. AT, UH, MD, CEG, SB, BG, and CG have verified the underlying data, AT performed the statistical analysis and prepared tables and figures. CG, CEG, and CS wrote the first manuscript draft. VRZ, VT, PAK, RRB, MM, APP, RS, AZ, JHB, AF, MS, HP, JCS, RT, GMK, JB, RvK, and STS contributed to interpretation of the results and critical revision of the manuscript. SB, KPB, CEG, CG, SDB, CA, DP, and BW implemented and coordinated the recruitment of study patients and biobank samples. TS, JT, DH, PDWG, JCS, ADC, CH, MP, DFS, LM, MCG, ASZ, AH, MG, NM, AR, FF, AT, JB, and AP contributed to the enrolment of study patients and data collection. All authors approved the final manuscript. CG is the guarantor for the study. The corresponding author attests that all listed authors meet authorship criteria and that no others meeting the criteria have been omitted.

### Copyright/ licence for publication

The corresponding author has the right to grant on behalf of all authors and does grant on behalf of all authors, a worldwide licence to the Publishers and its licensees in perpetuity, in all forms, formats and media (whether known now or created in the future), to i) publish, reproduce, distribute, display and store the Contribution, ii) translate the Contribution into other languages, create adaptations, reprints, include within collections and create summaries, extracts and/or, abstracts of the Contribution, iii) create any other derivative work(s) based on the Contribution, iv) to exploit all subsidiary rights in the Contribution, v) the inclusion of electronic links from the Contribution to third party material where-ever it may be located; and, vi) licence any third party to do any or all of the above.

---

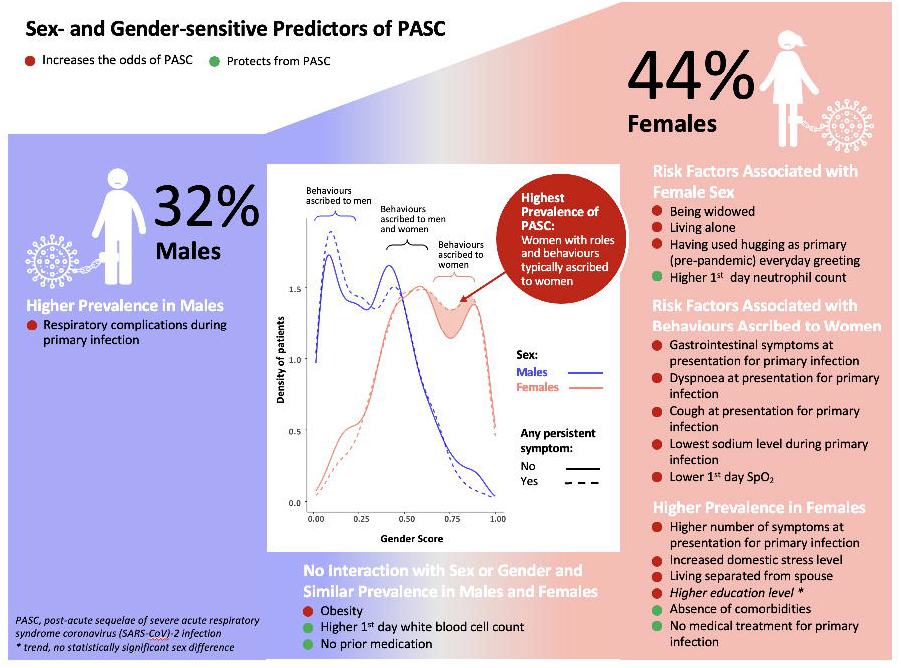

