## Supplementary material for "Understanding the Impact of Sociocultural Gender on Post-acute Sequelae of COVID-19: a Bayesian Approach": Supplemental Materials.docx

**Supplemental Material**

**Content**

**Supplemental Methods**

Study design and procedures

Participants versus non-participants

Follow-up questionnaire and definitions

Laboratory parameters

Construction of the Gender score

Statistical analyses

Statistical model in hospitalized patients

**Supplemental Results**

Patient baseline characteristics

Socioeconomic and behavioral characteristics

Acute COVID-19 disease course in overall population

Acute COVID-19 disease course in hospitalized patients

Laboratory parameters during primary infection in hospitalized patients

Post-COVID-19 sequelae

Sex-specific patients' characteristics

Interactions terms

Sex- and gender-specific determinants of PASC in the overall population

Sex- and gender-specific determinants of PASC in hospitalized patients

**References**

**Supplemental Figures**

**Supplemental Figure 1:** Flowchart depicting patient selection

**Supplemental Figure 2:** Gender-related questions included in the questionnaire

**Supplemental Figure 3:** Graphical illustration of statistical approach.

**Supplemental Figure 4:** Persistent symptoms reported at follow-up stratified by sex and	symptom.

**Supplemental Figure 5:** Visualization of the interaction between gender score and cough,	gastrointestinal symptoms, dyspnoea at presentation for acute COVID-19, and education	level.

**Supplemental Figure 6:** Visualization of the interaction between gender score and first day	SpO_2_, lowest sodium level, and first day neutrophil count during hospitalization for primary	infection

**Supplemental Figure 7:** Number of comorbidities, percentage of patients living separated or	divorced, percentage of patients being widowed or living alone, reported domestic stress,	cortisol level in	hospitalized patients by sex and persistence of symptoms.

**Supplemental Tables**

**Supplemental Table 1**: Acute COVID-19 disease characteristics I

**Supplemental Table 2:** Sociocultural- and economic variables

**Supplemental Table 3:** Acute COVID-19 disease characteristics II

**Supplemental Table 4:** Inflammatory markers and hormone levels during acute illness	**Supplemental Table 5:** Routine laboratory parameters during acute illness

**Supplemental Table 6:** Post-COVID-19 sequalae

**Supplemental Table 7:** Sex-specific patient characteristics

**Supplemental Methods**

**Study design and procedures**

Details on patient selection and recruitment are provided in Supplemental **Figure 1**.

Patients were recruited between February and December 2020 at one of four Swiss study sites including the University Hospital Basel, the University Hospital Zurich, the University Hospital Bern, and the Cantonal Hospital of Baden. In reporting this study, we have followed the STROBE guidelines for observational studies.(1) Both outpatients as well as hospitalized patients comprising normal ward, intermediate care (IMC), and ICU patients were included in our study. We deliberately allowed for inclusion of patients in additional observational studies (data of 16 intensive care unit [ICU] patients included in a previous observational study, PMID 32925314).

Study team members were trained to follow a standardized guideline for telephone interviews. All questionnaire data were collected through the Research Electronic Data Capture (REDCap) survey system to minimize missing inputs and allow for real-time data validation and quality control.(2, 3) Clinical data were obtained from electronic medical records and contained information about demographic characteristics (age, sex), cardiovascular risk factors including diabetes mellitus, hypertension, dyslipidaemia, family history of coronary artery disease, smoking, and obesity), symptoms and time of symptom onset, medication, pre-existing comorbidities, data on weight and height, laboratory parameters and disease severity of COVID-19 classified according to symptoms and necessity of in-hospital (normal ward, IMC or ICU) treatment. Items related to the sociocultural dimension gender were assessed by questionnaires as reported below.(4, 5) The study team entered and stored these data in the dedicated electronic database form REDCap which was hosted locally at the University Hospital Basel.

**Participants versus non-participants**

Compared to individuals not participating in our study-specific follow-up, patients who completed follow-up were older (median and interquartile range [IQR]: 42[30-57] vs 37[27-52] years, p<0.001), had more often hypertension (18.5% vs 15.3%, p=0.0013), were less often smokers (7.5% vs 11.3%, p<0.001), and suffered more often from chronic conditions (39.5% vs 35.4%, p=0.0013). No significant differences were observed for other cardiovascular risk factors (p=NS), pre-existing cardiovascular disease (11.1% vs 9.5%, p=0.052), the percentage of patients being hospitalized during primary infection (18.2% vs 15.5%, p=0.06), for female sex (45.8% vs 43.6%, p=0.095), and BMI (27[24-30] kg/m^2^ vs 27[24-31] kg/m^2^, p= 0.563.

**Follow-up questionnaire and definitions**

The follow-up questionnaire consisted of 58 questions for women and 51 questions for men and contained questions on socio-demographics, lifestyle variables, medical comorbidities, and risk factors, details on acute SARS-CoV-2 infection, current health status and symptoms, healthcare contacts since diagnosis, quality of life (QoL), gender-related and sex-specific parameters (**Supplemental Figure 2**).

Acute COVID-19 was defined as symptoms, consequences, and healthcare contacts within four weeks of first diagnosis. The disease severity during acute COVID-19 was characterized by (1) a three-category scale which consisted of the following categories: not admitted to hospital, admitted to normal ward, admitted to IMC or ICU. No pre-specified definition of PASC was applied and patients were asked to indicate when their symptoms were present and if/when symptoms had resolved at the time of follow-up. The overall QoL was assessed by using a five-category scale (“currently much better than before COVID-19”, “currently slightly better than before COVID-19”, “about the same as before COVID-19”, “currently slightly worse than before COVID-19”, “currently much worse than before COVID-19”). Domestic stress level was assessed by using a 10-category scale. Anxiety and depression were assessed by using the five-category scale of EQ-5D-5L instrument (“I am not anxious or depressed”, “ I am a little anxious or depressed”, “I am moderately anxious or depressed”, “I am very anxious or depressed”, “I am extremely anxious or depressed”).(6) To capture the longer-term effects of SARS-CoV-2 infection, we evaluated whether patients who were symptomatic in the acute phase had fully recovered compared to their normal health status before infection using a two-category scale (feeling “recovered and symptom free” or “not symptom free”.) The presence and type of any new or ongoing symptoms, that were not present prior to primary infection, was assessed using a comprehensive list of symptoms. Additionally, the research team reviewed and coded comments in free text fields for further new or ongoing symptoms not captured by the preconceived questionnaire. Further, patients were asked in the questionnaire to elaborate on their experience (if not captured by the preconceived questionnaire) in free text for the following areas: pre-existing comorbidities, medications prior to COVID-19 infection, and specific hormone treatments. Body mass index (BMI) was calculated from measured height and weight, and obesity was defined as BMI of 30kg/m^2^ or more. For both symptoms of PASC and symptoms of acute SARS-CoV-2 infection, it was differentiated between *non-specific* (=any) acute or persisting somatic symptoms as well as *specific* acute or persisting somatic symptoms of COVID-19. The latter comprised a cluster of somatic symptoms including dyspnea/reduced exercise performance, changes in smell and taste, which have previously been shown to be distinctive features of COVID-19 in both women and men.(7-11)

**Laboratory parameters**

Routine laboratory parameters such as white blood cells, platelet count, haemoglobin, high-sensitivity C-reactive protein, creatinine, sodium, liver transaminases, and cardiac troponin were immediately measured in fresh samples. Glomerular filtration rate (GFR) was calculated using the Modification of Diet in Renal Disease (MDRD) Study equation. Biobank samples were collected in standard blood collection tubes (Vacutainer™, Beckton Dickinson, Franklin Lakes, NewJersey, US) and stored in cryovials at –80 °C until final assessment. Testosterone, oestradiol, progesterone, and cortisol were analysed using electro-chemiluminescence-immunoassays (Elecsys®; Generation I for oestradiol, progesterone, and cortisol; Generation II for testosterone) on a cobas® 8000 e801 module (Roche Diagnostics, Rotkreuz, Switzerland). All assays were performed according to the manufacturers’ protocols and under strict internal and external quality control in the Department of Clinical Chemistry, University Hospital Zurich, Switzerland. The coefficient of variation (in %) and lower detection limit of these assays were as follows: testosterone with 2,2%, 2,9% and 0.087 nmol/L; oestradiol with 3,1%, 1,3% and 18.4 pmol/L; progesterone with 3,3%, 3,1%, and 0.159 nmol/L; cortisol with 3,4%, 3,5% and 1.5 nmol/L. In our cohort, samples of 3 patients were below the lower detection limit of oestradiol, 2 patients below the lower detection limit of testosterone, and 18 patients below the lower detection limit of progesterone.

**Construction of the Gender score**

Sex, which is defined as a person’s biological characteristics such as sex chromosomes, hormone concentrations, and sex organ physiology is distinct from gender, which refers to either social roles based on the sex of a person (“gender role”) or an individual’s identification of one's own gender based on an internal awareness (“gender identity”).(12) Both sex and gender are increasingly recognized as major determinants of health and disease outcomes,(13) however, the intertwining of these variables factoring into gender differences makes it difficult to differentiate between biological mechanisms and sex-specific behavior. In our study, we aimed to capture the two variables biological sex and gender by the study questionnaire. We assessed sex as a binary variable with the answering options “male” or “female”. The assessment of gender requires a complex approach as this variable consists of four interrelated dimensions (definition provided by the WHO(14) and the Women Health Research Network of the Canadian Institute of Health Research(15)) encompassing gender roles (e.g. child care), gender identity (a personal conception of oneself as man or woman), “gender relationships” (e.g. social support), and “institutionalized gender” (e.g. education level, personal income), which are experienced through the embodiment of norms, roles, values, and expectations, and expressed through ideas, behaviours, and attitudes. Accordingly, the challenge of assessing gender as a sociocultural construct lies in the fact that a large number of variables needs to be taken-into account, which often exceeds the capacities of a statistical approach in a study focusing on health outcomes. To reduce complexity and to maintain power of analysis it is desirable to reduce the number of variables and to express “gender” by a single variable, that can be entered into multivariate analysis similar to biological sex. We therefore applied a methodological approach from Pelletier et al. who constructed and validated a gender score as a continuous variable between zero (behaviours typically ascribed to men) to 1.00 (behaviours typically ascribed to women). This is based on a Canadian-Swiss questionnaire which includes a number of sociocultural variables that were "historically reported as being different in men and women”.(16, 17) The questionnaire was adapted for the German societal system and validated in a German cohort.(4, 18) The questionnaire was reduced to the seven items with the greatest predictive value for gender and adapted to the Swiss societal system. The resulting short gender questionnaire is provided in **Supplemental Figure 2** and contains seven questions on employment status, perceived social standing, housework responsibility status, education level, social support, domestic stress level as well as the Bem Sex-Role Inventory(19) (short version), a measure used to assess gender roles. These variables were included in a previously described logistic regression model using biological sex as the dependent variable.(16) The identified gender-related variables served as predictors to estimate the “probability of an individual being a woman” which was named gender score. The gender score ranged between 0.00 and 1.00 with higher values reflecting characteristics traditionally ascribed to women and lower values characteristics traditionally ascribed to men. Intermediate scores represent patients with an equal level of characteristics traditionally ascribed to women and men. Missing values were present in all gender-related variables, most prominently in questions being part of the Bem Sex-Role Inventory (2.87% missing), but missing values were below 2% in all other variables. In detail, out of 2927 patients 2654 (1238[92.4%] women, 1426[89.9%] men) answered the full set of gender-related questions required to calculate the gender score. The resulting gender score for our study population is depicted in **Figure 1C.** The mean gender score in our study population was 0.46±0.26 (range 0.00–1.00, **Supplemental Table 1**). As gender and sex usually overlap,(20) the gender score and biological sex were correlated in our study (Pearson r=0.52[0.49-0.55], p<0.001), which is consistent with data (r=0.62) reported previously by Pelletier et al.(16)

**Statistical analyses**

Bayesian models were used to estimate the probability of the primary and secondary study outcomes and the effect (log odds) of covariates in the overall population as well as in the subgroup of hospitalized patients (n=532).(21, 22) Bayesian modelling was employed because of the very low evidence-per-variable (EPV) measure of <10. Only cases with complete data were considered for statistical modelling, and no imputation was performed. Data were supplemented with publicly available records indicating prevalence and incidence of COVID-19 at the time of diagnosis of acute SARS-CoV-2 infection for each patient. The primary study outcome of persisting somatic symptoms was modelled using all available variables, including viral load for hospitalized patients, as well as interactions of each variable with biological (binary) sex and the gender score. The priors were set as follows: For the intercept, a normal distribution with mean 0 and standard deviation 1 was assumed to allow for a flexible baseline probability by having a weakly informative prior. A Laplace distribution with mean 0 and a spread estimated by the model was assumed for the independent variables, except for models with EPV <5, where a normal distribution with mean 0 and standard deviation of 0.00001 was assumed.(23) The Laplace prior was chosen to facilitate variable selection, as it would favor 0 as a coefficient for each independent variable. However, in cases where EPV was <5, this prior proved to be too restrictive and thus a weaker prior with normal distribution and standard deviation of 0.00001 was used. The full model was initially compared to a zero model which did not include any independent variables. Comparison of models were performed using the leave-one-out cross validation information criterion and the expected log pointwise predictive density (ELPD).(24) In brief, this measure can be used to estimate how well a model will predict future unknown data. If the full model significantly differed from the zero model, variable subsets(23) of the full model were created using the following procedure: Coefficient estimates intervals for the 10% to 90% highest posterior density were analysed at 10% interval steps. The range of vectors of coefficient estimates was considered across the sets of 10% to 90% highest posterior density values and variables were retained if their range was entirely above or below 0. Using this approach, successively smaller, more generalizable models were created with the same priors as the full model. All models were then compared based on ELPD and the most generalizable, smallest model was selected according to this metric. The model coefficients for the best models are displayed with the respective 90% credible interval. For comparison purposes, standard stepwise logistic regression with forward and backward stepwise selection using Akaike information criterion (AIC) was performed on the resulting best models where possible yet did not change the variable selection. The statistical approach is illustrated in **Supplemental Figure 3.**

**Statistical model in hospitalized patients (n=532):** With 57.9% of hospitalized patients (63.1% in women vs 55.2% in men) reporting persistent symptoms, this group represents by far the highest-risk group, where a timely risk prediction and initiation of preventive measures is urgently needed. In addition, given that the cohort of hospitalized patients is more homogenous than the outpatient cohort, the analysis of this subgroup provides important information that cannot be obtained from the overall cohort. Given the conflicting evidence regarding biological variables predicting PASC, we decided to include the maximum amount of laboratory parameters without pre-selection in our statistical model for hospitalized patients. This, however, led to the limitation of a low evidence per variable. On the other hand, this approach allows that amongst all variables (n=147), only the strongest predictor variables are selected by the model. Our results provide the basis for a better understanding of the etiology of PASC in women and men; however, the identified PASC risk conditions need to be confirmed in future studies.

**Supplemental Results**

**Patient baseline characteristics**

On average, men were older than women, and this age gap was particularly pronounced in patients with PASC (50[35-62] years vs 42[29-57] years, p<0.001, **Table 2**), while no significant sex differences were observed for BMI (27 [25-31] kg/m^2^ vs 27[23-31] kg/m^2^, p=0.345) **Table 2**). The most common risk condition in the overall cohort was hypertension (541 patients, 18.5%), followed by obesity (358 patients, 12.2%). Men more often had dyslipidemia, diabetes mellitus, or hypertension than women (p<0.01), independent of the presence or absence of PASC (**Table 2**). The percentage of male and female smokers amongst patients with PASC was similar (7.3% vs 7.4%, p=1.0). Cardiovascular disease including valvular heart disease, cardiomyopathies, or coronary artery disease, was present in 248 patients (8.5%) at the time of primary SARS-CoV-2 infection. Pre-existing cardiovascular disease was more common in men (p<0.001), while women suffered more often from autoimmune disorders, a sex difference that was more pronounced in patients without PASC (1.9% vs 4.8%, p=0.001) than in patients with PASC (2.4% vs 4.6%, p=0.064, **Table 2**). More men than women with PASC had cancer (8.3% vs 5.2%, p=0.049), while no sex difference in the prevalence of cancer was observed in patients without PASC (4.2% vs 5.0%, p=0.45). Women tended to have more often pre-existing mental disorders; however, this difference was not significant (p=NS, **Table 2**). Similarly, no statistically significant sex difference was seen for neurological or chronic pulmonary diseases (p=NS, **Table 2**), however, all these conditions were more common in patients with PASC as compared to those without PASC (**Table 2**). Individuals with PASC were more often on medications than individuals without PASC (**Table 2**). Independent of the presence or absence of PASC, men were more often on cardiovascular, lipid-lowering, anti-diabetic, and anti-thrombotic drugs than women (p<0.001).

**Acute COVID-19 disease course in overall population**

Out of 2927 individuals who were tested positive for SARS-CoV-2 and survived acute SARS-CoV-2 infection, 2395 (81.8%) remained outpatients, 532 (18.2%) were inpatients of whom 338 (11.6%) were admitted to a normal ward, and 194 (6.6%) were admitted to an IMC or ICU (**Supplemental Table 1**). Compared to women, men were more often inpatients (22.2% vs 13.4%, p<0.001**, Supplemental Table 1**). Individuals with PASC were more often hospitalized during acute illness as compared to individuals without PASC (28.2% vs 12.2%, p<0.001, **Supplemental Table 1**), independent of sex. The average number of reported symptoms during primary infection was higher in women as compared to men, and higher in individuals with PASC as compared to those without PASC (absence of PASC: 3.9±2.1 in men vs 4.6±2.2 in women, p<0.001; with PASC: 5.3±2.3 in men and 6.0±2.3 in women, p<0.001, **Supplemental Table 1**). This sex difference was less pronounced when only COVID-19 specific somatic symptoms including dyspnea/reduced exercise performance, dysgeusia, or anosmia were counted (**Supplemental Table 1**). A significant and positive correlation between the number of symptoms during primary infection and a higher gender score, indicating more feminine characteristics, was noted (Pearson r=0.12, p<0.001). Women reported more often gastrointestinal (GI) symptoms (24.3% vs 18.0%), changes in smell and taste, and fatigue (85.4% vs 77.9%) than men (p<0.02, **Supplemental Table 1**), while men presented more often with fever (64.2% vs 49.0%) than women (**Supplemental Table 1**).

**Socioeconomic and behavioral characteristics**

Most individuals in our study had undergone secondary education (39,3%) or had obtained a university/college degree (47.0%). Women with PASC had more often obtained a university or technical college degree than men in this group, however, these differences were not statistically significant (42.9% vs 37.6% in men, p=0.15, **Supplemental Table 2**). Conversely, women without PASC had less often obtained a university degree and more often undergone primary education than men in this group (p=0.027, **Supplemental Table 2**). Women were more often single parents than men, with the highest percentage of single parent women being observed in the PASC group (12.2% women vs 6.7% men, p<0.001, **Supplemental Table 2**). Also, women with PASC were more often divorced/separated (9.3% vs 7.3% in men, p=0.03), more often single (20.2% vs 15.6%), p=0.03 were more often widowed (4.8% vs 3.0%, p=0.03, **Supplemental Table 2** and **Supplemental Figure 4C**), and less often married/living in a partnership than men (65.5% vs 73.6%, p=0.029 for men vs women, **Supplemental Table 2**). These sex differences were also seen in patients without PASC but were less pronounced in this group (**Supplemental Figure 4D**). A higher percentage of individuals in the PASC group (12.3%) than in the non-PASC group (7.3%) were living without partner (widowed or divorced/separated) (**Supplemental Table 2**). Women reported higher stress levels at home than men with stress levels being highest in women with PASC (score 4.03±2.5 in women vs 3.22±2.11 in men, p<0.001, **Supplemental Table 2**, **Supplemental Figure 7E**). Conversely, during hospitalization, there was no evidence for a difference in cortisol levels between men and women or patients with and without PASC (**Supplemental Table 4**, **Supplemental Figure 7F**). Twenty percent of the study population were healthcare workers with significantly more women than men working in this sector (30.0% vs 12.0%, p<0.001 for men vs women, **Supplemental Table 2**). A greater percentage of women in our study reported earning the lowest income in a household than did male patients (40.7% of women vs 14.6% of men, p<0.001, **Supplemental Table 2**). Women were more often the main person responsible for household work (40.3% vs 14.2%, p<0.001). Women without PASC took over more responsibility than men for childcare/care of family members (score from 1 [minimum] to 6 [maximum]: 2.00±2.30 in women vs 1.67±2.12 in men, p=0.002), however, this sex difference was not seen in the PASC group (1.84±2.25 in women vs 1.72±2.07 in men, p=0.36, **Supplemental Table 2**). Women, independent of the presence or absence of PASC, self-reported higher female identity scores than men (score 1-7; 7=only feminine traits, 1=only masculine traits: 5.47±1.32 in women vs 2.37±1.48 in men without PASC, and 5.57±1.32 in women vs 2.26±1.39 in men with PASC; p<0.001). The overall gender score (range 0-1 with 1 reflecting traditionally feminine characteristics) was higher in women than in men, with differences between men and women being slightly more pronounced in individuals with PASC (without PASC: 0.34±0.22 in men vs 0.60±0.23 in women, p<0.001 and with PASC: 0.32±0.21 vs 0.62±0.22, p<0.001, **Supplemental Table 2**).

**Acute COVID-19 disease course in hospitalized patients**

In hospitalized patients, there was no evidence for sex differences in hemodynamic and respiratory parameters during primary infection in individuals who did not develop PASC, except for oxygen saturation, which was slightly lower in men as compared to women (**Supplemental Table 3**). Conversely, mean arterial pressure, PaO_2_/FiO_2_ ratio were lower, and respiratory rate was higher in hospitalized men who developed PASC as compared to women. Body temperature at presentation for primary infection was higher in men as compared to women, independent of PASC (p<0.02, **Supplemental Table 3**). Men who later developed PASC experienced more often than women, cardiac, renal, thromboembolic, or neurologic complications and received more often hemodynamic support and/or medical treatment of COVID-19 such as corticosteroids, remdesivir, chloroquine/hydroxychloroquine, tocilizumab, ritonavir/lopinavir (p<0.01 vs women, **Supplemental Table 3**). These sex differences were less pronounced or absent in patients who did not develop PASC (**Supplemental Table 3**).

**Laboratory parameters during primary infection in hospitalized patients**

Amongst hospitalized patients (n=532[18.2% of study population], 353[22.2%] men and 179[13.4%] women), there was strong evidence for sex differences in routine laboratory markers of inflammation with the maximum C-reactive protein level (median[IQR] 126.0[53.9-210.0] mg/L in men vs 54.0[17.0-147.8] mg/L in women, p<0.001), the maximum procalcitonin (0.26[0.12-1.03] µg/L in men vs 0.11[0.06-0.34] µg/L in women, p=0.008), the neutrophil:lymphocyte ratio (5.9[3.3-9.9] in men vs 3.6[2.3-7.3] in women, p=0.034), and the maximum ferritin levels (1132[712-2015] µg/L in men vs 459[191-1009] µg/L in women, p<0.001) being all higher in men (**Supplemental Table 4**). Similarly, a lower platelet count (163[127-221] G/L in men vs 202[158-241] G/L in women, p=0.019), higher D-Dimer levels (1.4[0.5-4.5] mg/L in men vs 0.7[0.4-2.3] mg/L in women, p=0.002), and higher levels of indicators of organ injury such as creatinine, liver transaminases, cardiac biomarkers as well as lactate were observed in men (**Supplemental Table 5)**. These laboratory parameters were consistently higher (inflammatory markers, D-Dimer, markers of organ injury, and lactate) or lower (platelet count) in patients who developed PASC as compared to patients who did not develop PASC (**Supplemental Table 4 and 5**). Notably, progesterone levels did not differ significantly between women and men, independent of PASC. The estradiol:testosterone ratio was lower in men as compared to women and this difference was most pronounced in patients who developed PASC (54.54[131.98] in men vs 422.25[958.17] in women, p=0.007, **Supplemental Table 4**).

**Post-COVID-19 sequelae**

The average number of specific and overall persistent symptoms at follow-up was higher in women as compared to men, however, this difference was statistically significant only for specific symptoms (specific symptoms: 1.3±1.0 in womenvs 1.2±1.0 in men, p<0.001, **Supplemental Table 6**), but not for any symptom (3.1±2.5 in women vs 3.0±2.3 in men, p=0.35, **Supplemental Table 6**). Individuals reporting a higher number of persistent symptoms more often suffered from anxiety or depression (Pearson r=0.308, p<0.001 in women and r=0.336, p<0.001 in men) and reported a higher stress level (Pearson r=0.226, p<0.001 in women and r=0.287, p<0.001 in men). In addition, the number of persistent symptoms showed a positive correlation with a higher gender score (=more feminine characteristics, Pearson r=0.059, p=0.002). The most frequent symptom at follow-up was reduced exercise tolerance and resilience in both sexes, which was reported by 43.8% of men and 41.5% of women followed by changes in smell and taste in 31.9% of women and 26.0 % of men) and shortness of breath in men (31.1 % of men and 29.7% of women, **Supplemental Figure 4**). Women were more often troubled with changes of smell and taste (22.5% vs 18.5%) as well as concentration deficits (28.0% vs 24.6%) than men, while men more often reported reduced exercise tolerance (43.8% vs 41.5%), shortness of breath (31.1% vs 29.7%) and chest pain (12.5% vs 11.2%) than women (**Supplemental Figure 4**). One-hundred-sixty-eight (5.8%) individuals (without PASC: 48[4.5%] men vs 28[3.7%] women, p=0.50; with PASC: 47[9.3%] men vs 45[7.7%] women, p=0.42) were re-hospitalized at least once for persistent symptoms or complications of COVID-19 **(Supplemental Table 6)**. Eight-hundred-sixty-three patients (29.5%) reported a worse QoL as compared to their pre-illness situation (p=NS for women vs men with and without PASC, **Supplemental Table 6**). The percentage of individuals reporting a worse QoL than before their illness was substantially higher in the PASC groups as compared to the non-PASC group (**Supplemental Table 6**). Men with PASC were more often dependent on help in everyday life as compared to women with PASC (7.3% vs 3.8% in women, p=0.015, **Supplemental Table 6**). No sex difference was observed in the percentage of individuals with PASC reporting pain or physical discomfort (p=0.54 for men vs women, **Supplemental Table 6**), however, women more often than men reported to be suffering from anxiety or depression since their primary infection. This sex difference was observed in the PASC group (p=0.025 for men vs women) as well as in the non-PASC group (p<0.001 for men vs women, **Supplemental Table 6**). A positive correlation between number of symptoms during acute infection and the presence of anxiety or depression was seen in both women (Pearson r=0.210, p<0.001) and men (r=0.191, p<0.001).

**Sex-specific patients' characteristics**

At the time of primary infection, 30.21% (n=404) of the female study population were postmenopausal. The percentage of postmenopausal women was higher in the PASC group as compared to those without PASC (33.5% vs 27.6%, p=0.018, **Supplemental Table 7**). Two-hundred-and-forty (17.9%) women had experienced pregnancy complications during a previous pregnancy with no difference in the incidence of previous pregnancy complications being seen between women with and without PASC (p=NS, **Supplemental Table 7**). Similarly, the number of pregnancies during lifetime was equally distributed between women with and without PASC (**Supplemental Table 7**). Forty women (3%) were pregnant at the time of primary infection (**Supplemental Table 7**). The number of pregnant women was higher in the group without PASC; however, this difference was not significant (3.7% vs 2.1%, p=0.11). There was no difference between the presence and absence of PASC in the number of women taking hormonal contraceptives (21.3% vs 20.1%, p=0.64) or postmenopausal hormone replacement therapies (2.0% vs 3.4%, p=0.14), while the number of women taking phytoestrogens was slightly higher for patients without PASC (p=0.043). There was also no difference between the absence and presence of- PASC in women who had undergone fertility treatments (0.7% vs 0.7%, p=1.0), with a history of polycystic ovary syndrome (4.3% vs 3.5%, p=0.55) or breast (2.4% vs 1.7%, p=0.52) or gynecological cancer (0.8% vs 0.3%, p=0.48). In men, the number of patients who had a history of prostate cancer was higher in the PASC group as compared to the group without PASC (2.4% vs 0.9%, p=0.04), however, no difference in the percentage of men receiving anti-androgenic treatment for prostate cancer at the time of primary infection was observed (0.2% vs 0.6%, p=0.39). More men in the PASC group as compared to the group without PASC were taking testosterone supplements at the time of primary infection (2.4% vs 0.9%, p=0.038, **Supplemental Table 7**).

**Interactions terms**

**Biological and sociocultural determinants of PASC in the overall population:**

**Figure 5** visualizes some of the interaction terms showing a strong association with PASC: Cough at presentation for acute COVID-19 showed a stronger association with the occurrence of PASC in individuals with a higher gender score (=more feminine characteristics). Conversely, in individuals with a lower gender score (=more masculine characteristics), cough at presentation for acute COVID-19 was associated with the absence of PASC (**Figure 5A**). Similar interactions and trends were observed for GI symptoms (**Figure 5B**), or dyspnea (**Figure 5C**) at presentation for acute COVID-19. A higher education level was associated with the occurrence of PASC, and women with PASC had more often obtained a university or technical college degree than men in this group, while an inverse trend was observed in individuals without PASC (**Supplemental Table 2**). **Figure 5D** visualizes these associations in relation to the gender score and depicts that, amongst individuals with a university of college degree (**lower panel**), more feminine characteristics were associated with PASC while more masculine characteristics were associated with the absence of PASC.

**Biological and sociocultural determinants of PASC in hospitalized patients**

Amongst hospitalized patients (n=532, 66.4% men and 33.6% women) typical feminine characteristics in combination with lower oxygen saturation at the first day of hospitalization were associated with the development of PASC (OR 1.60[1.30-1.96]) while being female and having a higher neutrophil count at the first day of hospitalization (OR 0.76[0.58-0.94]) or typical feminine characteristics in combination with lowest sodium level during primary infection (OR 0.74[0.64-0.84]) were negative predictors of PASC. Notably, as depicted in **Figure 3** some variables such as oxygen saturation, platelet count, or sodium level exerted opposite effects on the outcome measure when included in interaction with sex or gender score as compared to their effect as single variable. **Supplemental Figure 6** visualizes some of the above interactions by categorizing patients according to tertiles of the gender score or sex. Overall, individuals with PASC had lower SpO_2_ values at hospital admission for primary infection, however, this association was more evident in individuals with a lower gender score (=more masculine characteristics) than in individuals with a higher gender score (=feminine characteristics) (**Supplemental Figure 6A**). Similarly, individuals with PASC had lower sodium levels during hospital stay. However, this association was present in individuals with an intermediate and higher gender score (feminine and both feminine and masculine characteristics), but not in individuals with a low gender score (=masculine characteristics) (**Supplemental Figure 6B**). Finally, while a higher 1^st^ day neutrophil count seemed to be a protective factor for the occurrence of PASC in women, an opposite association was observed in men (**Supplemental Figure 6C**).

**Supplemental Figures**

**Supplemental Figure 1**
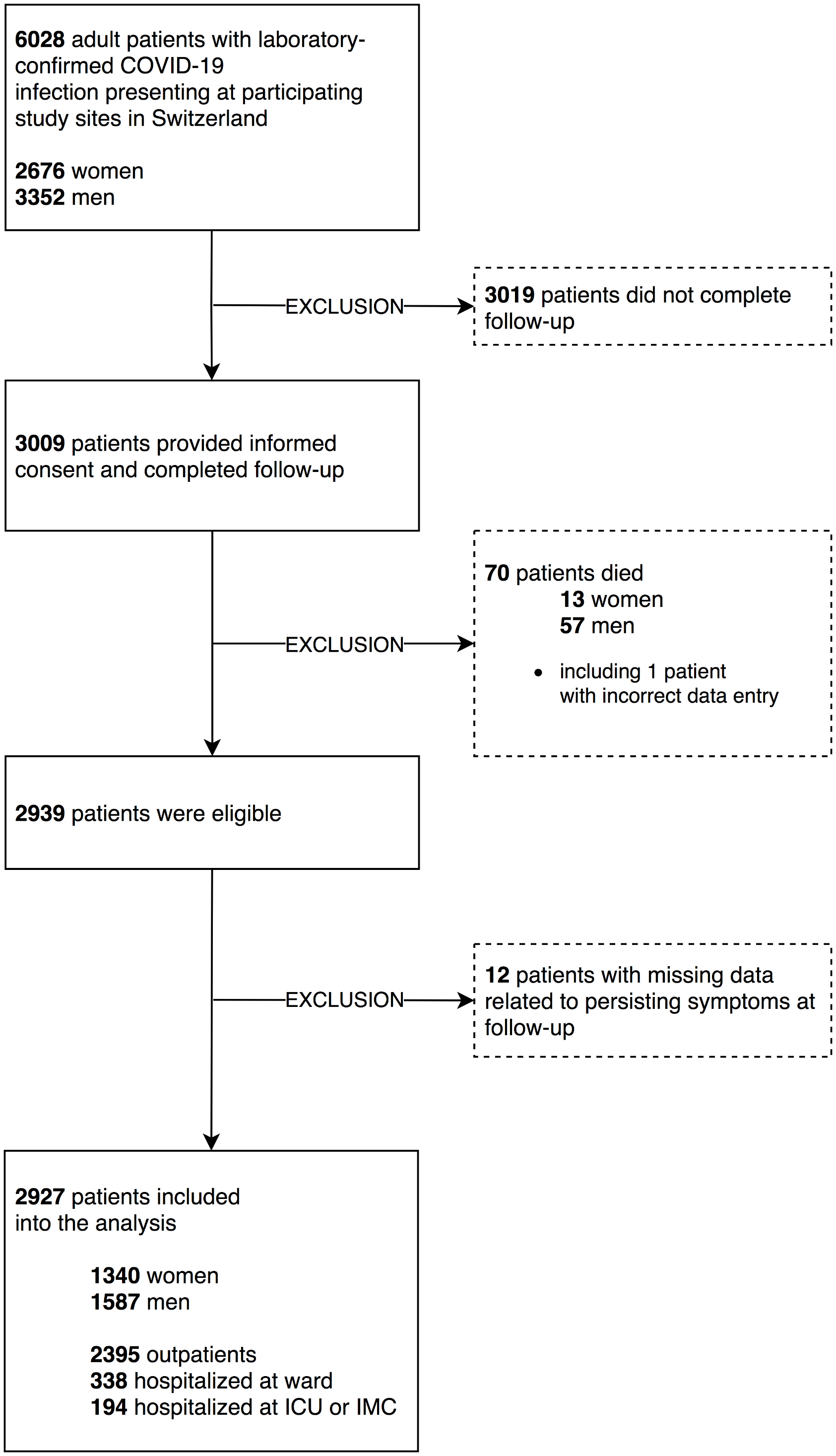

Flowchart depicting patient selection. ICU, intensive care unit; IMC, intermediate care unit.

**Supplemental Figure 2**

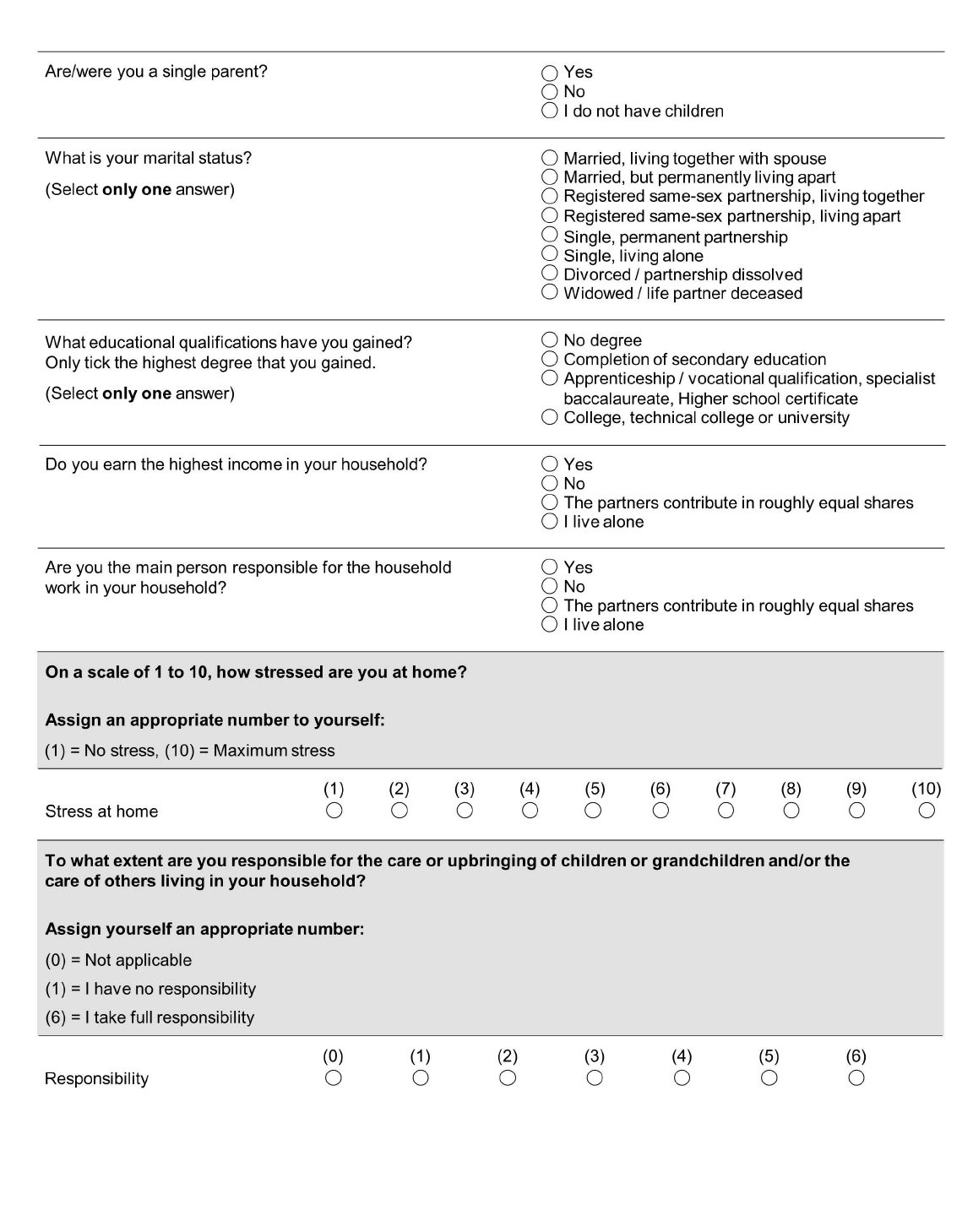

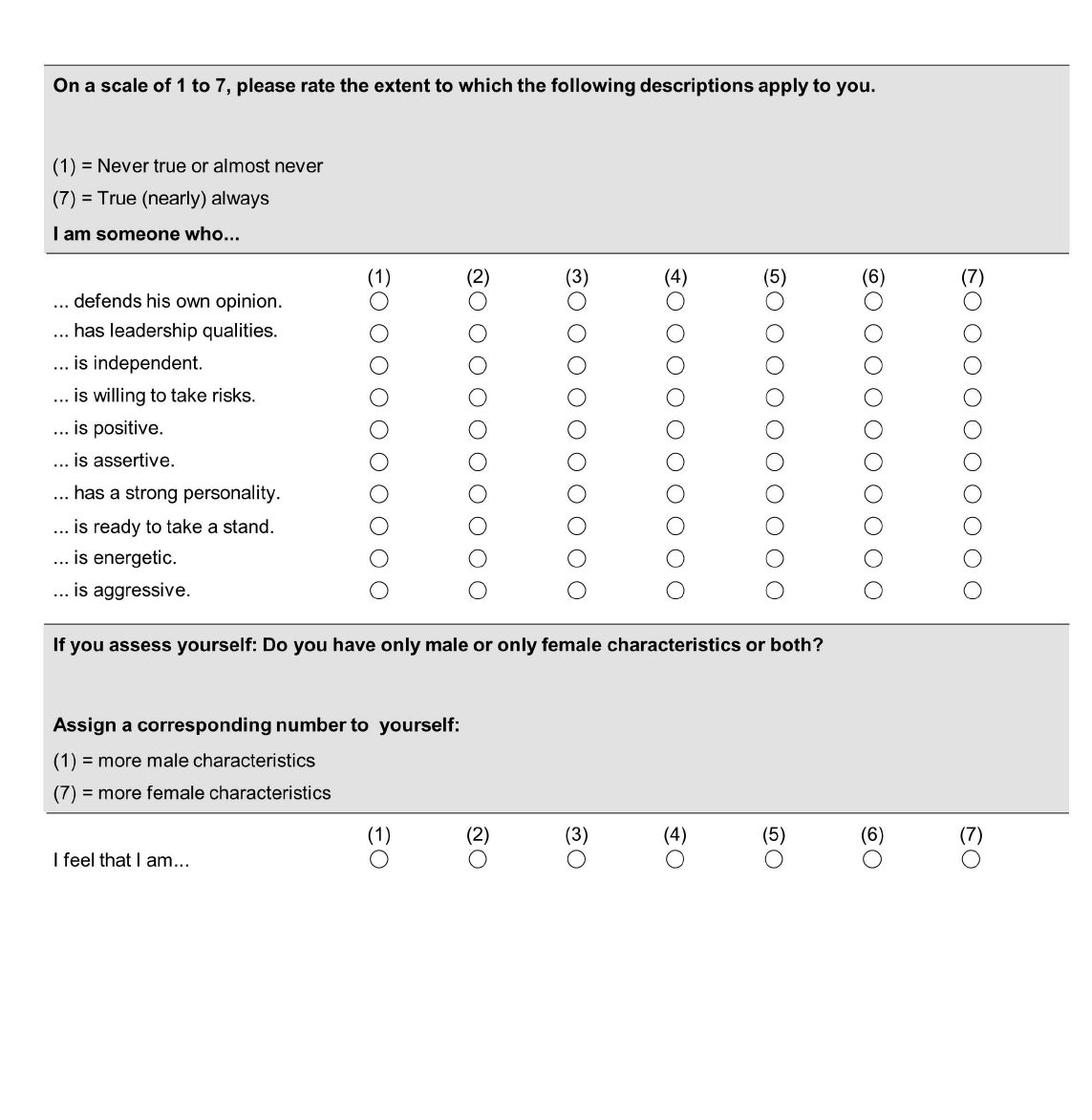

Gender-related questions included in the questionnaire.

**Supplemental Figure 3**

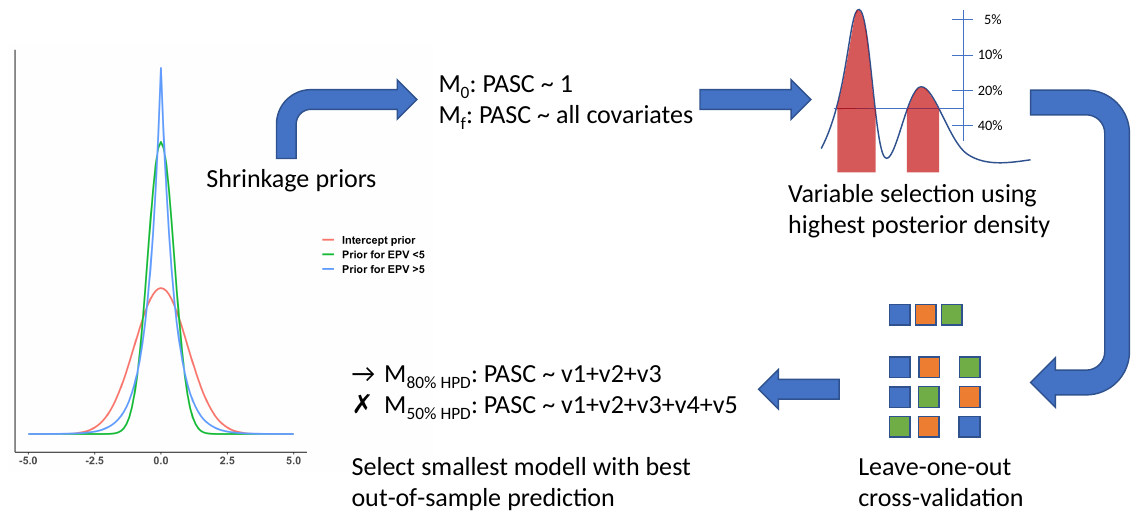

Graphical illustration of statistical approach. EPV, evidence-per-variable; HPD, highest posterior density; M, model; PASC, post-acute sequelae of SARS-CoV-2 infection.

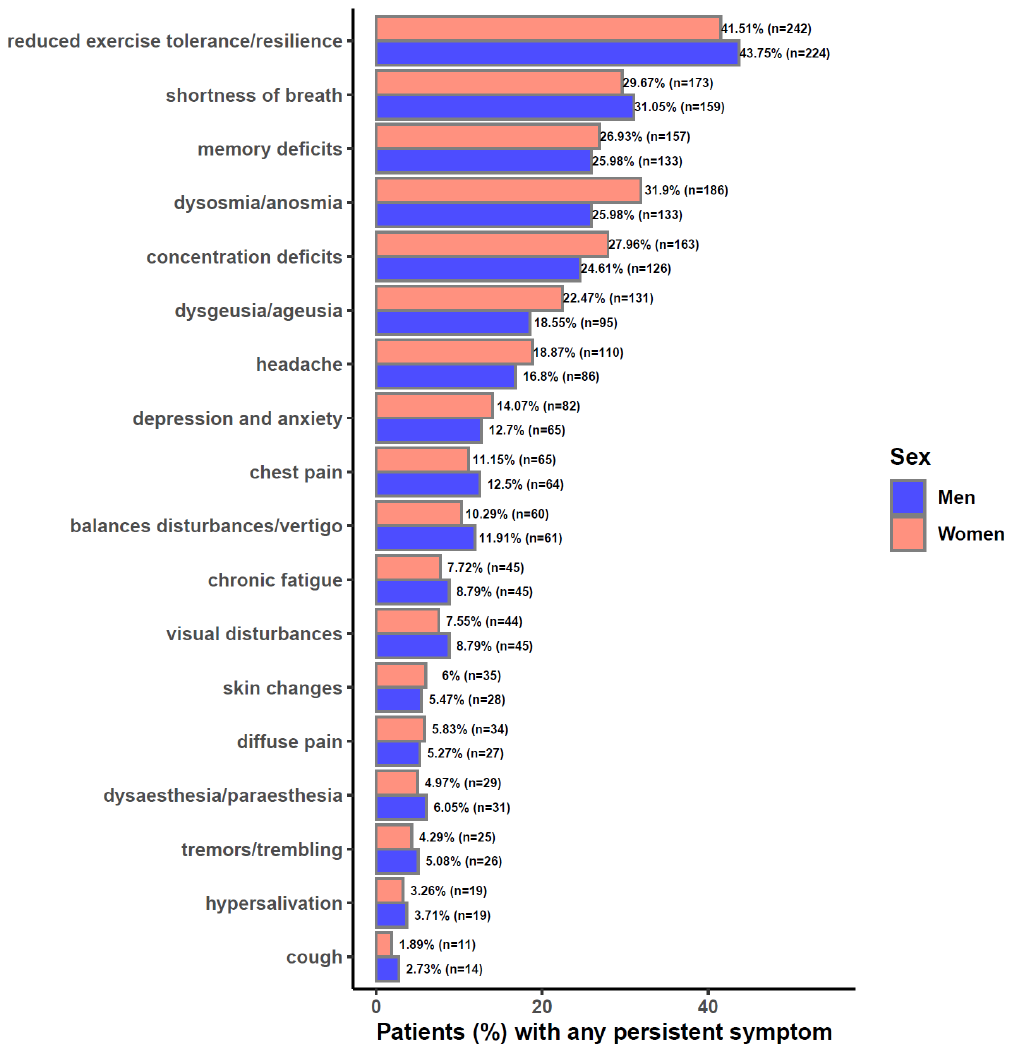
**Supplemental Figure 4**

Persistent symptoms reported at follow-up stratified by sex and symptom. Data are presented as percentage of patients (583 women and 508 men) reporting persistent symptoms.

**Supplemental Figure 5**

**A**

**B**

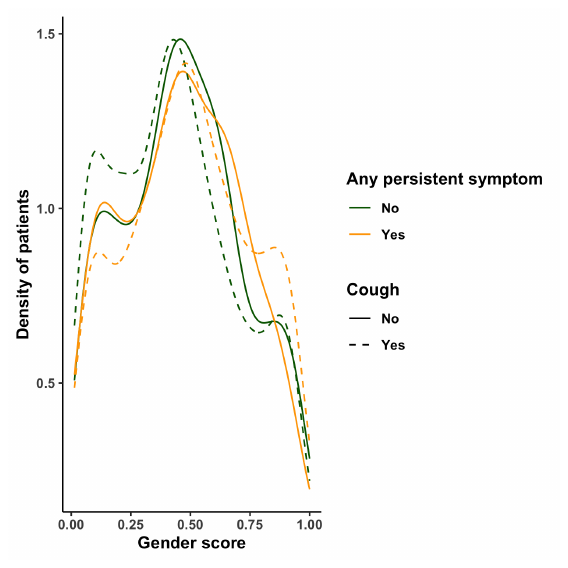

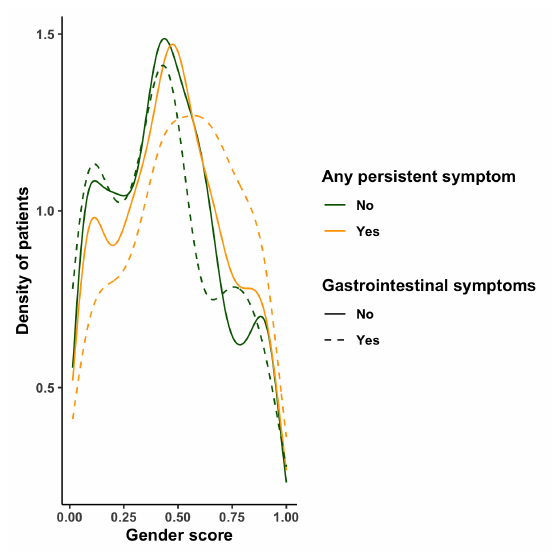

Masculine characteristics

Masculine characteristics

Feminine

characteristics

Feminine

characteristics

**C**

**D**

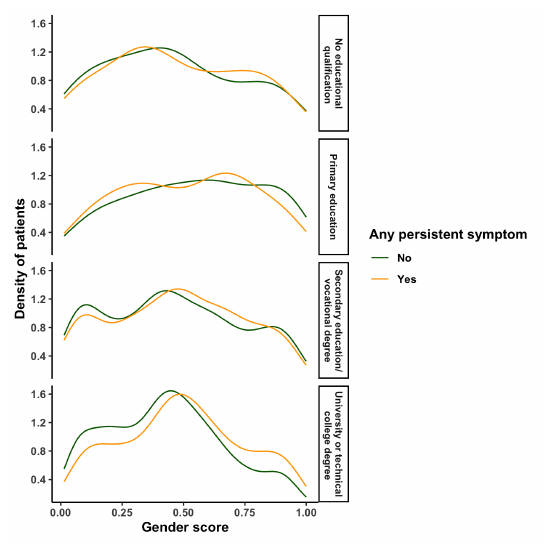

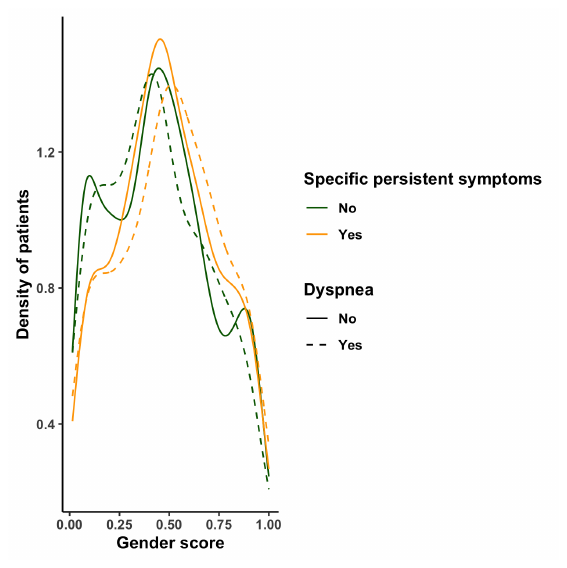

Masculine characteristics

Feminine

characteristics

Masculine characteristics

Feminine

characteristics

Visualization of the interaction between gender and **(A)** cough, **(B)** gastrointestinal symptoms, or **(C)** dyspnoea at presentation for acute COVID-19 as well as education level **(D)** for the primary endpoint of any persisting somatic symptom following SARS-CoV-2 infection.

**Supplemental Figure 6**

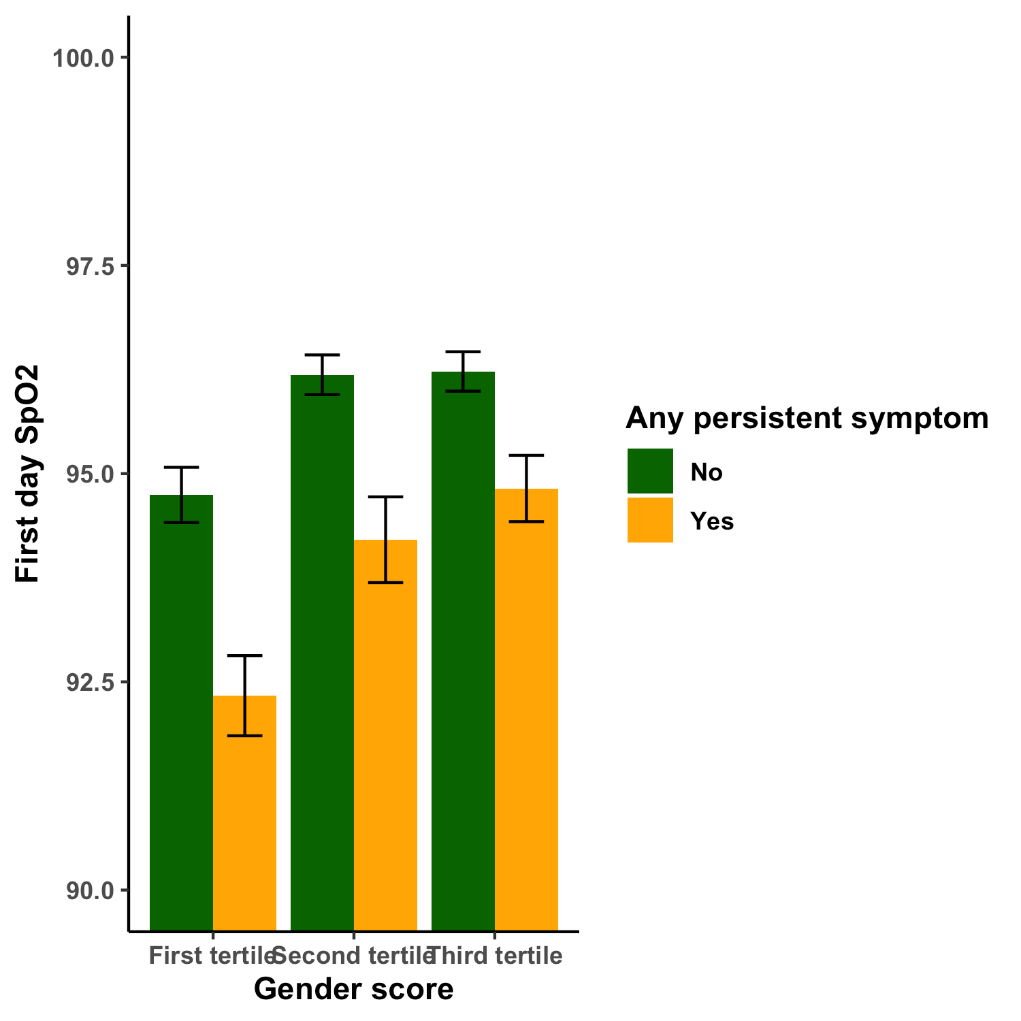

**A**

**B**

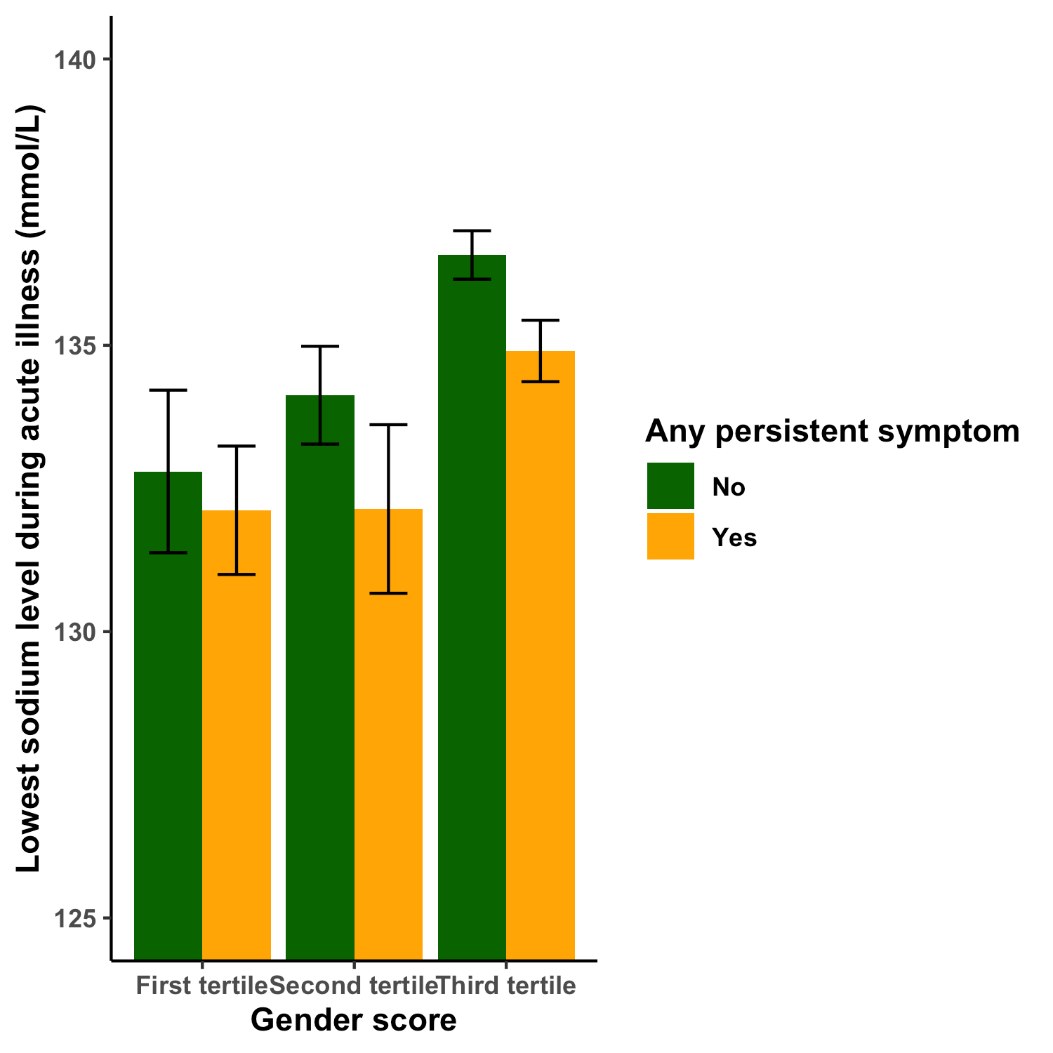

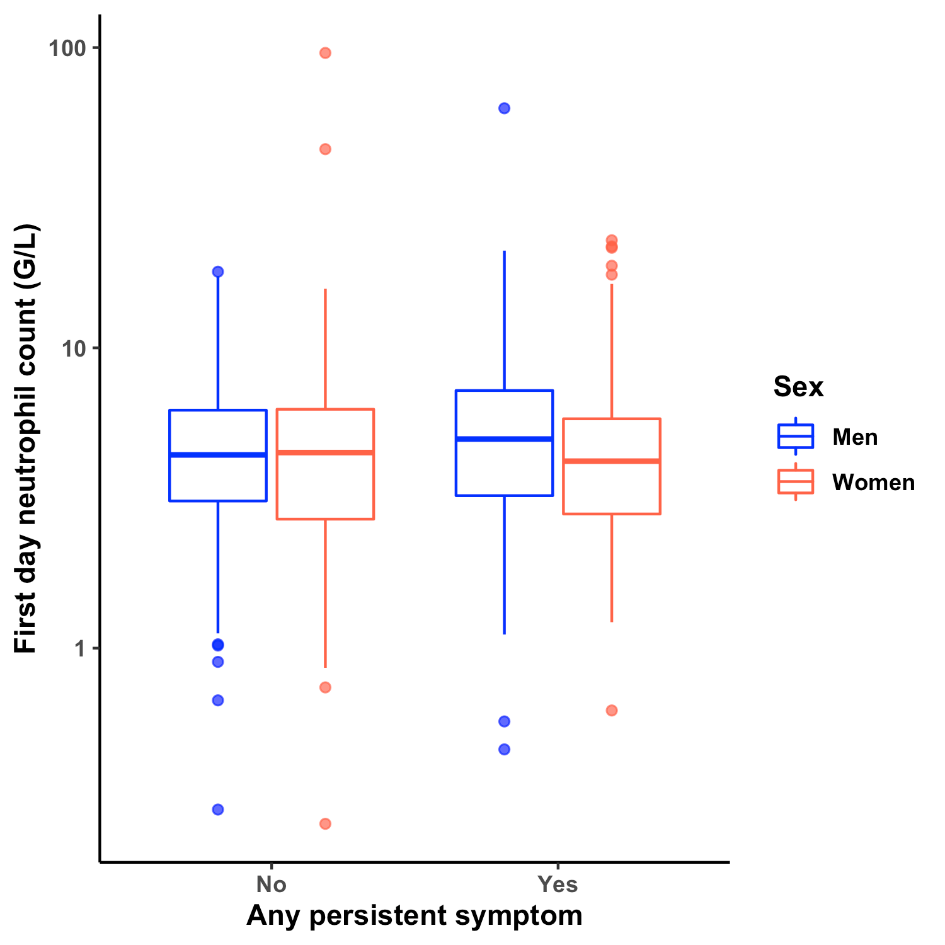

**C**

Visualization of the interaction between gender score (stratified by tertiles) and (A) First day SpO2 and (B) lowest sodium level during hospitalization for primary infection. (C) Boxplots of first day neutrophil count during primary infection by sex and PASC, illustrating the interaction between sex and PASC.

**Supplemental Figure 7**

**B**

**A**

**Number of comorbidities**

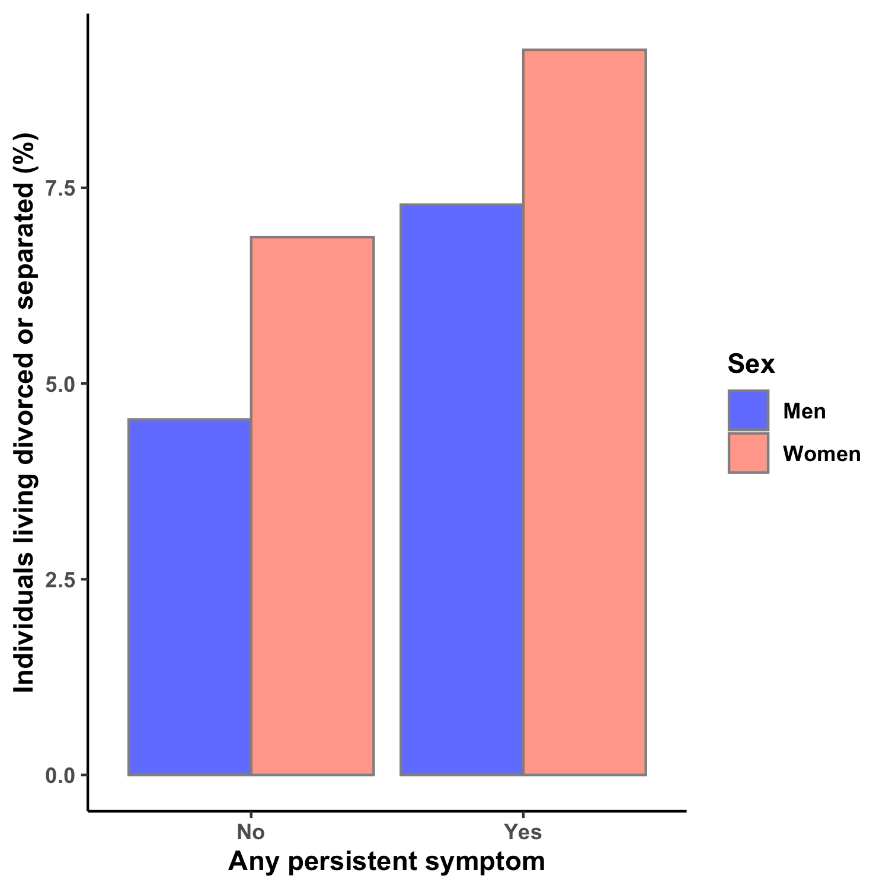

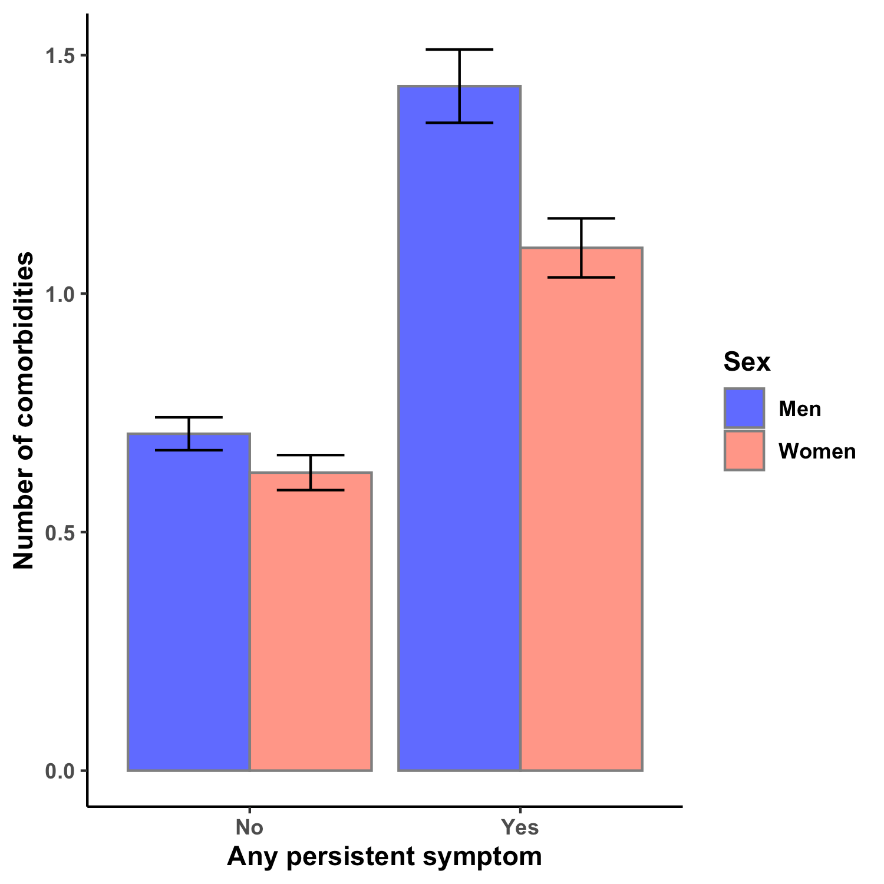

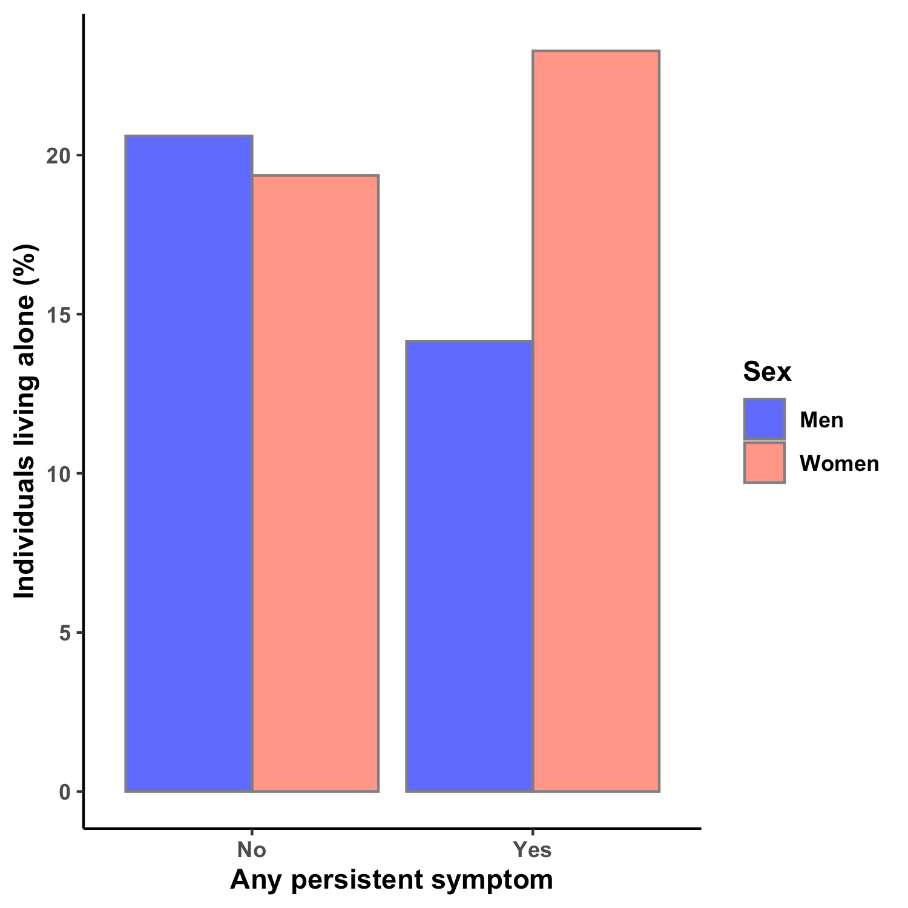

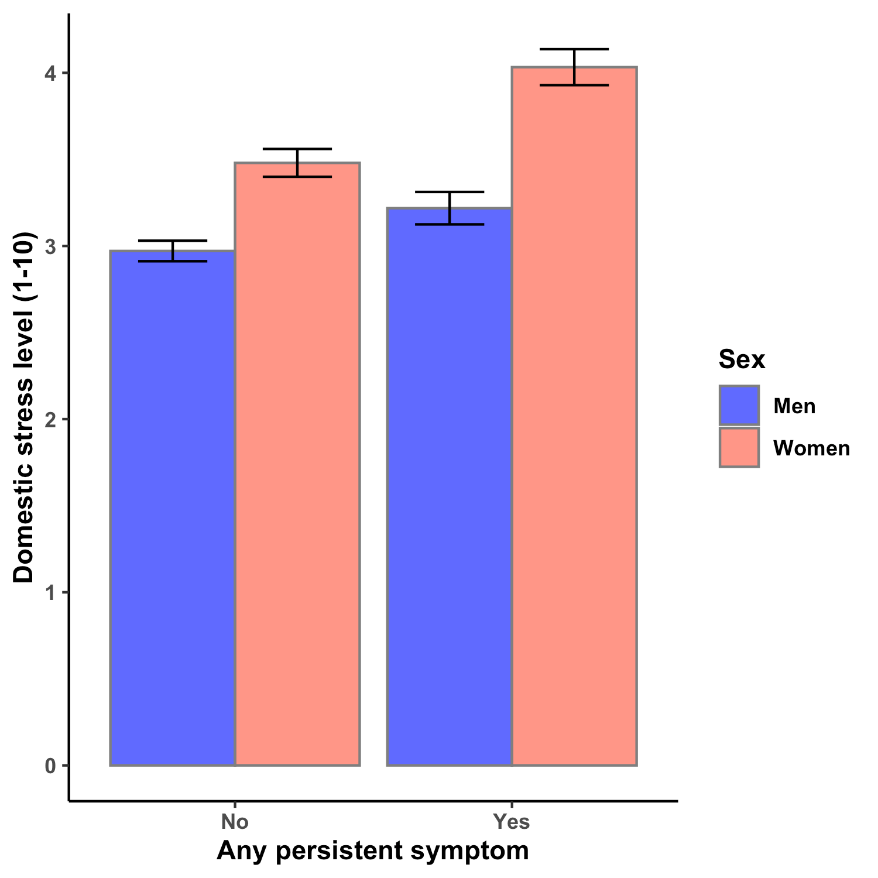

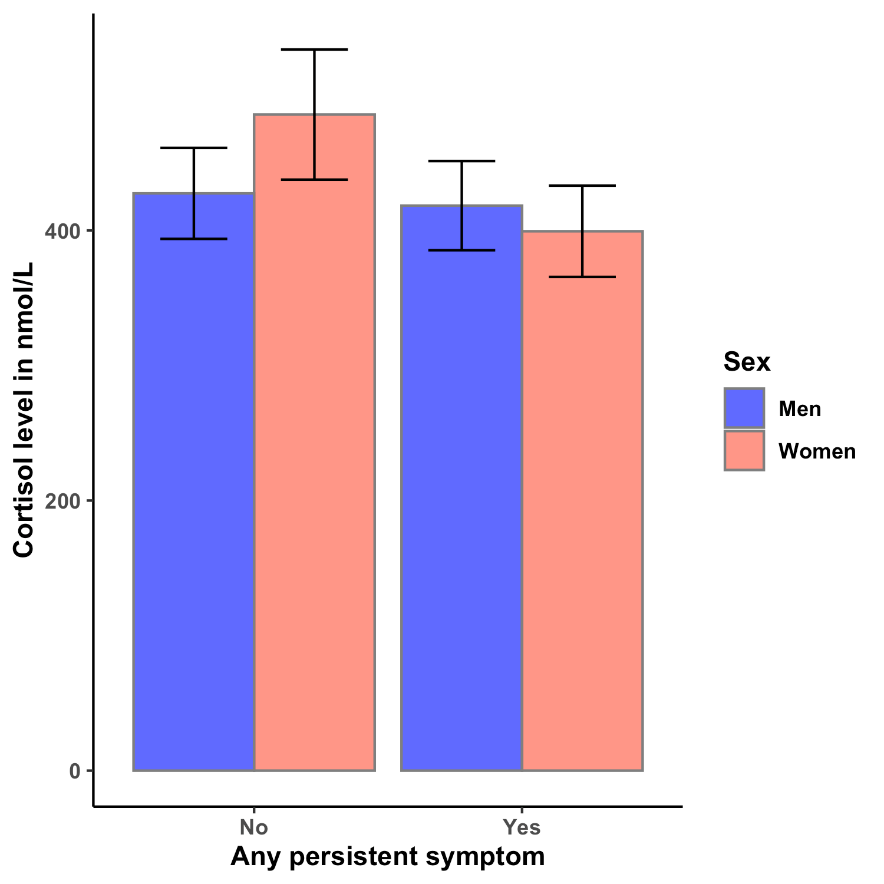

**C**

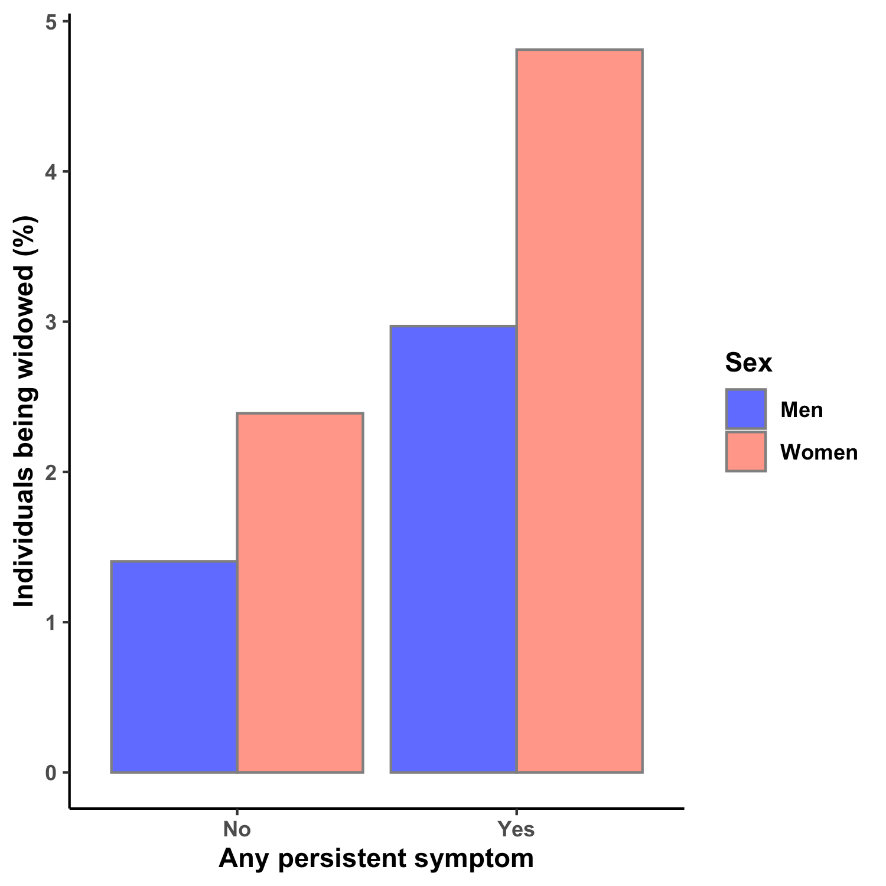

**D**

**E**

**F**

Number of comorbidities (means and standard error bars) **(A),** percentage of patients (total study population) living separated or divorced **(B)**, percentage of patients (total study population) being widowed **(C)** or living alone **(D)** by sex and persistence of symptoms. **(E)** Reported domestic stress level in the total study population (means and standard error bars, scale 0-10) by sex and persistence of symptoms. **(F)** Cortisol level in hospitalized patients (means and standard error bars) by sex and persistence of symptoms.
