## Supplemental Tables for "Understanding the Impact of Sociocultural Gender on Post-acute Sequelae of COVID-19: a Bayesian Approach"

| **Supplemental Table 1**<br>**Acute COVID-19 disease characteristics I** | **Overall (n=2927)** | **Absence of PASC** | | | **Presence of PASC** | | |
|---|---|---|---|---|---|---|---|
| | | **Males (n=1079)** | **Females (n=757)** | **p-value (men vs women)** | **Males (n=508)** | **Females (n=583)** | **p-value (men vs women)** |
| **Disease severity** | | | | | | | |
| Outpatient, n(%) | 2395 (81.8) | 921 (85.4) | 691 (91.3) | <0.001 | 313 (61.6) | 470 (80.6) | <0.001 |
| Normal ward hospitalization, n(%) | 338 (11.6) | 110 (10.2) | 52 (6.9) | | 104 (20.5) | 72 (12.4) | |
| Intermediate or intensive care unit hospitalization, n(%) | 194 (6.6) | 48 (4.5) | 14 (1.9) | | 91 (17.9) | 41 (7.0) | |
| **Symptoms at presentation for acute COVID-19** | | | | | | | |
| Number of **any** reported symptoms, mean(SD) | 4.8 (2.3) | 3.9 (2.1) | 4.6 (2.2) | <0.001 | 5.3 (2.3) | 6.0 (2.3) | <0.001 |
| Number of **specific** reported symptoms, mean(SD) | 2.3 (1.4) | 2.0 (1.3) | 2.1 (1.3) | 0.035 | 2.7 (1.3) | 2.9 (1.3) | 0.018 |
| Percentage of individuals reporting **specific** symptoms, n(%) | 2630 (89.9) | 922 (85.5) | 655 (86.5) | 0.559 | 488 (96.1) | 565 (96.9) | 0.550 |
| Gastrointestinal symptoms, n(%) | 571 (19.5) | 116 (10.8) | 136 (18.0) | <0.001 | 130 (25.6) | 189 (32.4) | 0.016 |
| Dyspnea, n(%) | 721 (24.6) | 150 (13.9) | 125 (16.5) | 0.140 | 203 (40.0) | 243 (41.7) | 0.607 |
| Cough, n(%) | 1394 (47.6) | 478 (44.3) | 308 (40.7) | 0.136 | 284 (55.9) | 324 (55.6) | 0.961 |
| Anosmia/dysosmia, n(%) | 1556 (53.2) | 458 (42.5) | 406 (53.6) | <0.001 | 287 (56.5) | 405 (69.5) | <0.001 |
| Ageusia/dysgeusia, n(%) | 1521 (52.0) | 441 (40.9) | 407 (53.8) | <0.001 | 287 (56.5) | 386 (66.2) | 0.001 |
| Fever, n(%) | 1550 (53.0) | 584 (54.1) | 333 (44.0) | <0.001 | 309 (60.8) | 324 (55.6) | 0.091 |
| Fatigue, n(%) | 2129 (72.7) | 683 (63.3) | 550 (72.7) | <0.001 | 401 (78.9) | 495 (84.9) | 0.013 |
| No symptoms, n(%) | 113 (3.9) | 64 (5.9) | 41 (5.4) | 0.714 | 4 (0.8) | 4 (0.7) | 1.0 |

Acute COVID-19 disease characteristics stratified by PASC (post-acute sequelae of SARS-CoV-2 infection) and sex. SD, standard deviation. P-values are reported for comparison between males and females. Comparisons were performed using t-test, non-parametric rank sum test, or chi-square test as appropriate. *only in hospitalized patients.

| **Supplemental Table 2:** **Sociocultural- and economic variables** | **Overall (n=2927)** | **Absence of PASC** | | | **Presence of PASC** | | |
|---|---|---|---|---|---|---|---|
| | | **Males (n=1079)** | **Females (n=757)** | **p-value (men vs women)** | **Males (n=508)** | **Females (n=583)** | **p-value (men vs women)** |
| Education | | | | 0.027 | | | 0.149 |
| • University or technical college degree, n(%) | 1375 (47.0) | 568 (52.6) | 366 (48.4) | | 191 (37.6) | 250 (42.9) | |
| • Secondary education or vocational degree, n(%) | 1151 (39.3) | 380 (35.2) | 287 (37.9) | | 243 (47.8) | 241 (41.3) | |
| • Primary education, n(%) | 202 (6.9) | 59 (5.5) | 64 (8.5) | | 35 (6.9) | 44 (7.6) | |
| • No educational qualification, n(%) | 182 (6.2) | 65 (6.0) | 38 (5.0) | | 33 (6.5) | 46 (7.9) | |
| Healthcare worker, n(%) | 577 (19.7) | 118 (10.9) | 223 (29.5) | <0.001 | 61 (12.0) | 175 (30.0) | <0.001 |
| Preferred greeting ritual | | | | <0.001 | | | <0.001 |
| • No physical contact, n(%) | 414 (14.1) | 166 (15.4) | 98 (13.0) | | 69 (13.6) | 81 (13.9) | |
| • Handshake/handclap, n(%) | 1176 (40.2) | 591 (54.8) | 166 (22.0) | | 288 (56.7) | 131 (22.5) | |
| • Cheek kiss, n(%) | 376 (12.9) | 68 (6.3) | 146 (19.3) | | 44 (8.7) | 118 (20.2) | |
| • Hug/Embrace, n(%) | 738 (25.2) | 180 (16.7) | 273 (36.1) | | 69 (13.6) | 216 (37.1) | |
| Marital status | | | | 0.0125 | | | 0.029 |
| • Divorced/separated, n(%) | 192 (6.6) | 49 (4.5) | 52 (6.9) | | 37 (7.3) | 54 (9.3) | |
| • Married/partnership, n(%) | 2046 (69.9) | 750 (69.5) | 540 (71.3) | | 374 (73.6) | 382 (65.5) | |
| • Single, n(%) | 594 (20.3) | 254 (23.5) | 143 (18.9) | | 79 (15.6) | 118 (20.2) | |
| • Widowed, n(%) | 76 (2.6) | 15 (1.4) | 18 (2.4) | | 15 (3.0) | 28 (4.8) | |
| Parenthood | | | | 0.003 | | | <0.001 |
| • Single-parent family, n(%) | 227 (7.8) | 57 (5.3) | 65 (8.6) | | 34 (6.7) | 71 (12.2) | |
| • Two-parent family, n(%) | 1682 (57.5) | 652 (60.4) | 411 (54.3) | | 323 (63.6) | 296 (50.8) | |
| • No children, n(%) | 988 (33.8) | 358 (33.2) | 276 (36.5) | | 145 (28.5) | 209 (35.9) | |
| Income | | | | <0.001 | | | <0.001 |
| • Earns highest income in household, n(%) | 1048 (35.8) | 528 (48.9) | 151 (20.0) | | 260 (51.5) | 109 (18.7) | |
| • Equal between partners, n(%) | 452 (15.4) | 140 (13.0) | 122 (16.1) | | 91 (17.9) | 99 (17.0) | |
| • Earns lowest income in household, n(%) | 826 (28.2) | 180 (16.7) | 335 (44.3) | | 74 (14.6) | 237 (40.7) | |
| • Lives alone, n(%) | 571 (19.6) | 220 (20.4) | 146 (19.3) | | 70 (13.8) | 135 (23.2) | |
| Main person responsible for household work, n(%) | | | | <0.001 | | | <0.001 |
| • No, n(%) | 695 (23.7) | 342 (31.7) | 110 (14.6) | | 181 (35.6) | 62 (10.6) | |
| • Equal distribution between partners, n(%) | 975 (33.3) | 370 (34.3) | 239 (31.6) | | 192 (37.8) | 174 (29.9) | |
| • Yes, n(%) | 771 (26.3) | 179 (16.6) | 285 (37.7) | | 72 (14.2) | 235 (40.3) | |
| • Lives alone, n(%) | 457 (15.6) | 177 (16.4) | 117 (15.5) | | 54 (10.6) | 109 (18.7) | |
| Main responsibility for childcare/care of family members (min 1-max 6), mean (SD) | 1.79 (2.2) | 1.67 (2.1) | 2.00 (2.3) | 0.002 | 1.72 (2.1) | 1.84 (2.3) | 0.364 |
| Average domestic stress level (scale 1-10, 10=maximum), mean (SD) | 3.36 (2.2) | 2.97 (1.9) | 3.48 (2.2) | <0.001 | 3.22 (2.1) | 4.03 (2.5) | <0.001 |
| Self-assessment of gender identity (scale 1-7, 7=only feminine traits, 1=only masculine traits), mean (SD) | 3.79 (2.1) | 2.37 (1.5) | 5.47 (1.3) | <0.001 | 2.26 (1.4) | 5.57 (1.3) | <0.001 |
| Overall Gender score, mean (SD) | 0.46 (0.3) | 0.34 (0.2) | 0.60 (0.2) | <0.001 | 0.32 (0.2) | 0.62 (0.2) | <0.001 |

Socioeconomic characteristics of the **total study population** stratified by post-COVID-19 syndrome and sex. PASC, post-acute sequelae of SARS-CoV-2 infection; SD, standard deviation. P-values are reported for comparison between males and females. Comparisons were performed using t-test, non-parametric rank sum test, or chi-square test as appropriate.

| **Supplemental Table 3:** **Acute COVID-19 disease characteristics II** | **Overall (n=2927)** | **Absence of PASC** | | | **Presence of PASC** | | |
|---|---|---|---|---|---|---|---|
| | | **Males (n=1079)** | **Females (n=757)** | **p-value (men vs women)** | **Males (n=508)** | **Females (n=583)** | **p-value (men vs women)** |
| **Clinical parameters at presentation for acute COVID-19** | | | | | | | |
| CURB-65, mean(SD)* | 1.6 (1.3) | 1.5 (1.3) | 1.3 (1.1) | 0.430 | 1.8 (1.3) | 1.6 (1.4) | 0.329 |
| Physician's disease severity index (PDSI), mean(SD) | 1.6 (0.9) | 1.5 (0.9) | 1.4 (0.7) | 0.397 | 1.8 (1.1) | 1.7 (1.0) | 0.372 |
| Mean arterial pressure (MAP) (mmHg), median(IQR) | 88 (77-100) | 91 (80-102) | 88 (78-100) | 0.216 | 85 (73-95) | 89 (78-101) | 0.002 |
| Heart rate (HR) (beats/minute), median(IQR) | 86 (74-99) | 84 (72-100) | 85 (75-99) | 0.542 | 88 (75-101) | 87 (75-98) | 0.223 |
| Respiratory rate (breaths/minute), median(IQR) | 20 (16-25) | 20 (16-25) | 19 (16-22) | 0.157 | 22 (18-29) | 20 (17-25) | 0.003 |
| P/F ratio, (mmHg), median(IQR)* | 297 (211-343) | 311 (233-343) | 311 (257-397) | 0.240 | 272 (152-325) | 297 (204-361) | 0.016 |
| Oxygen saturation ($SpO_2$) (%), median(IQR) | 96 (93-98) | 96 (93-98) | 97 (96-98) | <0.001 | 93 (89-97) | 97 (94-98) | <0.001 |
| Body temperature (°C), median(IQR) | 37.2 (36.7-38.0) | 37.2 (36.5-38.0) | 37.0 (36.6-37.7) | 0.014 | 37.5 (36.8-38.5) | 37.3 (36.7-37.9) | <0.001 |
| **Disease course of acute COVID-19** | | | | | | | |
| Respiratory complications, n(%)* | 434 (14.8) | 124 (11.5) | 40 (5.3) | <0.001 | 175 (34.5) | 95 (16.3) | <0.001 |
| Invasive ventilation, n(%)* | 124 (4.2) | 23 (2.1) | 2 (0.3) | 0.014 | 66 (13.0) | 33 (5.7) | 0.156 |
| Hemodynamic support, n(%)* | 127 (4.3) | 26 (2.4) | 6 (0.8) | 0.015 | 66 (13.0) | 29 (5.0) | <0.001 |
| Cardiac complications, n(%)* | 76 (2.6) | 10 (0.9) | 7 (0.9) | 1.0 | 43 (8.5) | 16 (2.7) | <0.001 |
| Renal complications, n(%)* | 94 (3.21) | 22 (2.0) | 5 (0.7) | 0.027 | 48 (9.5) | 19 (3.3) | <0.001 |
| Thromboembolic complications, n(%)* | 52 (1.8) | 10 (0.9) | 1 (0.1) | 0.062 | 31 (6.1) | 10 (1.7) | <0.001 |
| Neurological complications, n(%)* | 86 (2.9) | 16 (1.5) | 5 (0.7) | 0.159 | 50 (9.9) | 15 (2.6) | <0.001 |
| Medical treatment of acute COVID-19* | | | | | | | |
| - corticosteroids, n(%) | 207 (7.1) | 56 (5.2) | 15 (2.0) | <0.001 | 93 (18.3) | 43 (7.4) | <0.001 |
| - remdesivir, n(%) | 124 (4.2) | 39 (3.6) | 10 (1.3) | 0.004 | 53 (10.4) | 22 (3.8) | <0.001 |
| - chloroquine/hydroxychloroquine, n(%) | 123 (4.2) | 30 (2.8) | 13 (1.7) | 0.185 | 56 (11.0) | 24 (4.1) | <0.001 |
| - tocilizumab, n(%) | 45 (1.5) | 9 (0.8) | 0 (0.0) | <0.001 | 25 (4.9) | 11 (1.9) | 0.009 |
| - ritonavir/lopinavir, n(%) | 77 (2.6) | 23 (2.1) | 8 (1.1) | 0.115 | 31 (6.1) | 15 (2.6) | 0.006 |

Acute COVID-19 disease characteristics stratified by PASC (post-acute sequelae of SARS-CoV-2 infection) and sex. CURB-65: Confusion, Urea nitrogen, Respiratory rate, Blood pressure, 65 years of age and older; IQR, interquartile range; P/F ratio, $PaO_2/FiO_2$ ratio; SD, standard deviation. P-values are reported for comparison between males and females. Comparisons were performed using t-test, non-parametric rank sum test, or chi-square test as appropriate. *only in hospitalized patients.

| **Supplemental Table 4:** **Inflammatory markers and hormone levels during acute illness (hospitalized patients)** | **Overall (n=532)** | **Absence of PASC** | | | **Presence of PASC** | | |
|---|---|---|---|---|---|---|---|
| | | **Males (n=158)** | **Females (n=66)** | **p-value (men vs women)** | **Males (n=195)** | **Females (n=113)** | **p-value (men vs women)** |
| Highest virus load (copies/mL), median(IQR) | 74200 (7000-2660220) | 44500 (7000-2707300) | 10100 (5100-350000) | 0.285 | 97100 (7300-2819070) | 195000 (18950-2036325) | 0.233 |
| Highest white blood cell count (G/L), median(IQR) | 8.5 (6.0-12.8) | 7.9 (5.9-11.4) | 7.6 (5.3-10.6) | 0.097 | 9.9 (6.7-14.8) | 8.1 (5.9-12.7) | 0.492 |
| Lowest white blood cell count (G/L), median(IQR) | 5.1 (3.9-6.7) | 5.0 (3.8-6.5) | 5.6 (4.3-7.8) | 0.132 | 4.9 (3.8-6,7) | 5.1 (4.0-6.7) | 0.101 |
| 1st day white blood cell count (G/L), median(IQR) | 6.5 (4.7-8.6) | 6.1 (4.7-8.8) | 6.6 (4.7-8.4) | 0.628 | 6.7 (4.9-9.1) | 6.3 (4.4-8.1) | 0.462 |
| Neutrophil count (G/L), median(IQR) | 6.1 (4.0-10.0) | 5.3 (3.8-8.8) | 5.1 (3.7-7.8) | 0.101 | 7.8 (5.0-12.4) | 5.3 (3.6-9.2) | 0.012 |
| Neutrophil count (%), median(IQR) | 78.5 (68.4-87.0) | 76.9 (68.1-85.6) | 75.3 (63.0-82.2) | 0.047 | 83.8 (74.9-89.2) | 73.0 (65.0-84.1) | <0.001 |
| 1st day neutrophil count (G/L), median(IQR) | 4.4 (3.0-6.5) | 4.4 (3.1-6.2) | 4.5 (2.7-6.2) | 0.034 | 5.0 (3.2-7.2) | 4.2 (2.8-5.8) | 0.260 |
| Lowest lymphocyte count (G/L), median(IQR) | 0.8 (0.5-1.1) | 0.8 (0.5-1.1) | 1.0 (0.6-1.3) | 0.869 | 0.6 (0.4-0.9) | 0.9 (0.6-1.4) | 0.039 |
| Lowest lymphocyte count (%), median(IQR) | 12.6 (6.5-20.7) | 13.7 (7.6-19.8) | 14.7 (8.8-24.5) | 0.269 | 8.5 (4.8-14.8) | 17.8 (8.4-26.0) | 0.001 |
| 1st day lymphocyte count (G/L), median(IQR) | 0.9 (0.6-1.4) | 1.0 (0.6-1.3) | 1.1 (0.8-1.6) | 0.989 | 0.8 (0.5-1.1) | 1.1 (0.7-1.5) | 0.068 |
| Neutrophil:lymphocyte ratio, median(IQR) | 4.4 (2.5-8.5) | 4.6 (2.7-7.3) | 3.6 (2.0-6.6) | 0.263 | 5.9 (3.3-9.9) | 3.6 (2.3-7.3) | 0.034 |
| CRP (mg/L), median(IQR) | 84 (25-166) | 79 (22-161) | 39 (8-101) | 0.004 | 126 (54-210) | 54 (17-148) | <0.001 |
| Procalcitonin (µg/L), median(IQR) | 0.17 (0.08-0.52) | 0.15 (0.09-0.43) | 0.08 (0.06-0.23) | 0.452 | 0.26 (0.12-1.03) | 0.11 (0.06-0.34) | 0.008 |
| Ferritin (µg/L), median(IQR) | 841 (433-1721) | 1015 (485-1908) | 483 (223-915) | <0.001 | 1132 (712-2015) | 459 (191-1009) | <0.001 |
| Interleukin 6 (IL-6, ŋg/L), median(IQR)* | 103 (37-373) | 88 (27-301) | 31 (12-50) | 0.119 | 133 (55-421) | 102 (36-692) | 0.111 |
| 1st day cortisol, (nmol/L), mean (SD) | 423.56 (293.08) | 427.49 (261.13) | 485.85 (268.26) | 0.325 | 418.43 (326.89) | 399.43 (280.85) | 0.688 |
| 1st day estradiol (E2), (pmol/L), mean (SD) | 126.01 (146.37) | 91.14 (32.25) | 114.6 (154.26) | 0.432 | 116.52 (97.71) | 175.76 (230.28) | 0.054 |
| 1st day testosterone, (nmol/L), mean (SD) | 3.99 (5.09) | 6.31 (4.7) | 1.06 (1.64) | <0.001 | 5.64 (6.09) | 0.81 (1.11) | <0.001 |
| 1st day progesterone, (nmol/L), mean (SD)* | 0.78 (1.31) | 0.42 (0.36) | 0.61 (0.56) | 0.504 | 1.02 (1.65) | 0.86 (1.53) | 0.705 |
| Estradiol:testosterone ratio, mean (SD) | 150.68 (513.78) | 28 (29.71) | 191.61 (336.59) | 0.033 | 54.54 (131.98) | 422.25 (958.17) | 0.007 |

Inflammatory markers and hormone levels in **hospitalized patients**, stratified by PASC (post-acute sequelae of SARS-CoV-2 infection) and sex. CRP, C-reactive protein; IQR, interquartile range; LDH, lactate dehydrogenase; MDMR, modification of diet in renal disease; SD, standard deviation. P-values are reported for comparison between males and females. Comparisons were performed using t-test, non-parametric rank sum test, or chi-square test as appropriate. *Data only available in < 200 patients.

| **Supplemental Table 5:**<br>**Routine laboratory parameters during acute illness (hospitalized patients)** | **Overall (n=532)** | **Absence of PASC** | | | **Presence of PASC** | | |
|---|---|---|---|---|---|---|---|
| | | **Males (n=158)** | **Females (n=66)** | **p-value (men vs women)** | **Males (n=195)** | **Females (n=113)** | **p-value (men vs women)** |
| Lowest hemoglobin (g/L), median(IQR) | 122 (105-134) | 129 (115-140) | 123 (108-133) | 0.055 | 118 (98-133) | 118 (101-131) | 0.599 |
| Lowest platelet count (G/L), median(IQR) | 181 (136-236) | 171 (130-224) | 216 (160-279) | <0.001 | 163 (127-221) | 202 (158-241) | 0.019 |
| 1st day platelet count (G/L), median(IQR) | 203 (152-262) | 191 (140-241) | 229 (175-281) | 0.002 | 188 (143-250) | 218 (171-275) | 0.024 |
| Highest fibrinogen (g/L), median(IQR)* | 5.8 (4.2-7.7) | 5.1 (3.8-7.1) | 4.4 (3.3-5.9) | 0.369 | 6.3 (5.0-8.2) | 5.4 (4.6-7.7) | 0.159 |
| Highest D-Dimer (mg/L), median(IQR) | 0.9 (0.4-2.4) | 0.7 (0.4-1.4) | 0.6 (0.4-1.1) | 0.970 | 1.4 (0.5-4.5) | 0.7 (0.4-2.3) | 0.002 |
| Neuron-specific enolase (NSE, µg/L), median(IQR)* | 29.4 (22.5-38.8) | 29.1 (22.07-36.3) | 38.3 (36.3-40.4) | 0.820 | 28.1 (22.2-40.4) | 26.8 (21.7-33.0) | 0.892 |
| Highest ASAT (U/L), median(IQR) | 47 (31-89) | 48 (30-77) | 37 (24-48) | 0.034 | 60 (39-118) | 41 (27-83) | 0.217 |
| Highest ALAT (U/L), median(IQR) | 50 (27-100) | 52 (29-92) | 31 (18-55) | <0.001 | 74 (33-132) | 38 (23-85) | 0.003 |
| Highest bilirubin (µmol/L), median(IQR) | 10.0 (7.0-15.0) | 10.1 (7.2-15.2) | 7.6 (4.0-10.8) | <0.001 | 11.0 (8.4-17.0) | 8.1 (6.0-12.7) | 0.025 |
| 1st day bilirubin (µmol/L), median(IQR) | 8.0 (5.7-10.6) | 8.2 (7.0-11.9) | 6.0 (3.7-9.5) | <0.001 | 8.9 (6.6-11.8) | 7.0 (5.0-9.0) | <0.001 |
| Lowest albumin (g/L), median(IQR) | 32 (26-37) | 32 (28-38) | 34 (28-38) | 0.894 | 30 (23-35) | 33 (28-38) | 0.003 |
| Highest LDH (U/L), median(IQR) | 374 (263-568) | 346 (251-517) | 298 (231-412) | 0.027 | 449 (329-675) | 347 (235-527) | 0.054 |
| Highest creatinine (µmol/L), median(IQR) | 89 (72-110) | 94 (78-109) | 68 (58-82) | <0.001 | 103 (83-135) | 75 (62-97) | <0.001 |
| Lowest GRF (MDRD, mL/min/1.73m$^2$), median(IQR) | 71 (55-92) | 75 (61-93) | 88 (61-104) | 0.014 | 63 (46-87) | 67 (54-91) | 0.037 |
| Highest CK (U/L), median(IQR) | 118 (65-253 | 126 (72-233) | 77 (50-117) | 0.013 | 158 (91-498) | 77 (55-166) | 0.014 |
| Highest troponin T (ηg/L), median(IQR) | 15 (8-36) | 15 (8-27) | 13 (5-23) | 0.400 | 21 (10-54) | 10 (6-26) | 0.020 |
| Highest lactate (mmol/L), median(IQR) | 1.5 (1.1-2.3) | 1.5 (1.1-2.2) | 1.5 (0.9-1.9) | 0.995 | 1.7 (1.2-2.6) | 1.5 (1.0-2.2) | 0.027 |
| Lowest sodium (mmol/L), median(IQR) | 135 (132-138) | 136 (132-138) | 137 (134-139) | 0.002 | 134 (131-136) | 136 (134-138) | <0.001 |

Routine laboratory parameters in **hospitalized patients**, stratified by PASC (post-acute sequelae of SARS-CoV-2 infection) and sex. ALAT, alanine aminotransferase; ASAT, aspartate aminotransferase; CK, creatine kinase; GFR, glomerular filtration rate; IQR, interquartile range; LDH, lactate dehydrogenase; MDMR, modification of diet in renal disease; SD, standard deviation. P-values are reported for comparison between males and females. Comparisons were performed using t-test, non-parametric rank sum test, or chi-square test as appropriate.

| **Supplemental Table 6:** **Post-COVID-19 sequalae** | **Overall (n=2927)** | **Absence of PASC** | | | **Presence of PASC** | | |
|---|---|---|---|---|---|---|---|
| | | **Males (n=1079)** | **Females (n=757)** | **p-value (men vs women)** | **Males (n=508)** | **Females (n=583)** | **p-value (men vs women)** |
| Follow-up time, months, median (IQR) | 6 (5-9) | 6 (5-8) | 6 (5-9) | 0.142 | 6 (5-10) | 6 (5-0) | 0.808 |
| Number of **all** persisting symptoms, mean (SD) | 1.2 (2.1) | 0 | 0 | - | 3.0 (2.3) | 3.1 (2.5) | 0.354 |
| Number of **specific** persisting symptoms, mean (SD) | 0.5 (0.8) | 0 | 0 | - | 1.2 (1.0) | 1.3 (1.0) | 0.001 |
| Percentage of individuals reporting **specific** persisting symptoms, n(%) | 844 (28.8) | 0 | 0 | - | 380 (74.8) | 464 (79.6) | 0.070 |
| Hospital readmission, n(%) | 168 (5.7) | 48 (4.5) | 28 (3.7) | 0.494 | 47 (9.3) | 45 (7.7) | 0.417 |
| Quality of life (QoL) | | | | 0.822 | | | 0.346 |
| - Similar QoL than before COVID-19, n(%) | 1764 (60.3) | 815 (75.5) | 573 (75.7) | | 158 (31.1) | 218 (37.4) | |
| - Much better QoL than before COVID-19, n(%) | 115 (3.9) | 49 (4.5) | 29 (3.8) | | 18 (3.6) | 19 (3.3) | |
| - Much worse QoL than before COVID-19, n(%) | 138 (4.7) | 15 (1.4) | 10 (1.3) | | 54 (10.6) | 59 (10.1) | |
| - Slightly better QoL than before COVID-19, n(%) | 158 (5.4) | 72 (6.7) | 45 (5.9) | | 20 (3.9) | 21 (3.6) | |
| - Slightly worse QoL than before COVID-19, n(%) | 725 (24.8) | 119 (11.0) | 93 (12.3) | | 251 (49.4) | 262 (44.9) | |
| Every day-life | | | | 0.649 | | | 0.015 |
| - Independent, n(%) | 2780 (95.0) | 1034 (95.8) | 724 (95.6) | | 465 (91.5) | 557 (95.5) | |
| - Dependent on assistance, n(%) | 112 (3.9) | 29 (2.7) | 24 (3.2) | | 37 (7.3) | 22 (3.8) | |
| Individuals reporting pain/physical discomfort since COVID-19 disease | | | | 0.161 | | | 0.542 |
| - No pain/physical discomfort, n(%) | 1968 (67.2) | 951 (88.2) | 635 (83.9) | | 184 (36.2) | 198 (34.0) | |
| - Minimal pain/physical discomfort, n(%) | 579 (19.8) | 99 (9.2) | 93 (12.3) | | 180 (35.4) | 207 (35.5) | |
| - Moderate pain/physical discomfort, n(%) | 259 (8.9) | 16 (1.5) | 17 (2.3) | | 97 (19.1) | 129 (22.1) | |
| - Severe pain/physical discomfort, n(%) | 77 (2.6) | 8 (0.7) | 6 (0.8) | | 27 (5.3) | 36 (6.2) | |
| - Extreme pain/physical discomfort, n(%) | 15 (0.5) | 1 (0.1) | 1 (0.1) | | 8 (1.6) | 5 (0.9) | |
| Anxiety/depression since COVID-19 disease | | | | <0.001 | | | 0.025 |
| - No anxiety/depression since COVID-19 disease, n(%) | 2007 (68.6) | 862 (79.9) | 553 (73.1) | | 300 (56.1) | 292 (50.1) | |
| - Little anxiety/depression since COVID-19 disease, n(%) | 593 (20.3) | 166 (15.4) | 146 (19.3) | | 118 (23.2) | 163 (28.0) | |
| - Moderate anxiety/depression since COVID-19 disease, n(%) | 224 (7.7) | 39 (3.6) | 44 (5.8) | | 53 (10.4) | 88 (15.3) | |
| - Substantial anxiety/depression since COVID-19 disease, n(%) | 67 (2.3) | 4 (0.4) | 10 (1.3) | | 23 (4.5) | 30 (5.2) | |
| - Extreme anxiety/depression since COVID-19 disease, n(%) | 13 (0.4) | 5 (0.5) | 0 (0.0) | | 4 (0.8) | 4 (0.7) | |

Characteristics of post-acute COVID-19 sequalae of the **total study population** stratified by post-COVID syndrome and sex. IQR, interquartile range; PASC, post-acute sequelae of SARS-CoV-2 infection; SD, standard deviation. P-values are reported for comparison between males and females. Comparisons were performed using t-test, non-parametric rank sum test, or chi-square test as appropriate.

| **Supplemental Table 7:** **Sex-specific patient characteristics** | **Females** | | | | **Males** | | | |
|---|---|---|---|---|---|---|---|---|
| | **Overall women (n=1340)** | **No PASC (n=757)** | **PASC (n=583)** | **p-value** | **Overall men (n=1587)** | **No PASC (n=1079)** | **PASC (n=508)** | **p-value** |
| Menopause, n(%) | 404 (30.2) | 209 (27.6) | 195 (33.5) | 0.018 | - | - | - | - |
| Polycystic ovary syndrome, n(%) | 52 (3.9) | 32 (4.2) | 20 (3.4) | 0.550 | - | - | - | - |
| Pregnancy complications | | | | | - | - | - | - |
| - None, n(%) | 464 (34.6) | 267 (35.3) | 197 (33.8) | 0.612 | | | | |
| - Gestational diabetes, n(%) | 35 (2.6) | 18 (2.4) | 17 (2.9) | 0.660 | | | | |
| - Gestational hypertension, n(%) | 18 (1.3) | 8 (1.1) | 10 (1.7) | 0.425 | | | | |
| - Miscarriage, n(%) | 138 (10.3) | 71 (9.4) | 67 (11.5) | 0.242 | | | | |
| - Preterm delivery, n(%) | 7 (0.5) | 3 (0.4) | 4 (0.7) | 0.728 | | | | |
| - Preeclampsia-/eclampsia, n(%) | 8 (0.6) | 2 (0.3) | 6 (1.0) | 0.149 | | | | |
| - Other, n(%) | 34 (2.5) | 16 (2.1) | 18 (3.1) | 0.343 | | | | |
| Pregnancy at time of infection, n(%) | 40 (3.0) | 28 (3.7) | 12 (2.1) | 0.110 | - | - | - | - |
| Number of pregnancies during lifetime | | | | | - | - | - | - |
| - 0, n(%) | 616 (46.0) | 361 (47.7) | 255 (43.7) | 0.167 | | | | |
| - 1, n(%) | 196 (14.6) | 113 (14.9) | 83 (14.2) | 0.782 | | | | |
| - 2, n(%) | 264 (19.7) | 154 (20.3) | 110 (18.9) | 0.546 | | | | |
| - 3, n(%) | 140 (10.5) | 74 (9.8) | 66 (11.3) | 0.408 | | | | |
| - 4, n(%) | 50 (3.7) | 22 (2.9) | 28 (4.8) | 0.095 | | | | |
| - ≥5, n(%) | 37 (2.8) | 15 (2.0) | 22 (3.8) | 0.069 | | | | |
| Hormonal contraceptives, n(%) | 278 (20.8) | 161 (21.3) | 117 (20.1) | 0.639 | - | - | - | - |
| Postmenopausal hormone replacement, n(%) | 35 (2.6) | 15 (2.0) | 20 (3.4) | 0.140 | - | - | - | - |
| Fertility treatments, n(%) | 9 (0.7) | 5 (0.7) | 4 (0.7) | 1.000 | - | - | - | - |
| Intake of phytoestrogens | | | | 0.043 | | | | 0.124 |
| - Never, n(%) | 513 (38.3) | 269 (35.5) | 244 (41.9) | | 672 (42.3) | 438 (40.6) | 234 (46.1) | |
| - Rarely, n(%) | 523 (39.0) | 320 (42.3) | 203 (34.8) | | 657 (41.4) | 468 (43.4) | 189 (37.2) | |
| - Weekly, n(%) | 220 (16.4) | 124 (16.4) | 96 (16.5) | | 200 (12.6) | 134 (12.4) | 66 (13.0) | |
| - Daily, n(%) | 69 (5.2) | 38 (5.0) | 31 (5.3) | | 33 (2.1) | 23 (2.1) | 10 (2.0) | |
| Cancer | | | | | | | | |
| - Breast cancer, n(%) | 28 (2.1) | 18 (2.4) | 10 (1.7) | 0.517 | 0 (0.0) | 0 (0.0) | 0 (0.0) | - |
| - Gynaecologic cancer, n(%) | 8 (0.6) | 6 (0.8) | 2 (0.3) | 0.483 | - | - | - | - |
| - Prostate cancer, n(%) | - | - | - | - | 22 (1.4) | 10 (0.9) | 12 (2.4) | 0.040 |
| - Testicular cancer, n(%) | - | - | - | - | 5 (0.3) | 3 (0.3) | 2 (0.4) | 1.0 |
| Current anti-estrogen therapy for breast/gynaecologic cancer, n(%) | 17 (1.3) | 9 (1.2) | 8 (1.4) | 0.959 | - | - | - | - |
| Current anti-androgenic treatment for prostate cancer, n(%) | - | - | - | - | 5 (0.3) | 2 (0.2) | 3 (0.6) | 0.388 |
| Regular testosterone administration, n(%) | 0 (0.0) | 0 (0.0) | 0 (0.0) | - | 22 (1.4) | 10 (0.9) | 12 (2.4) | 0.038 |

Characteristics of post-acute COVID-19 sequalae of the **total study population** stratified by post-COVID syndrome and sex. IQR, interquartile range; PASC, post-acute sequelae of SARS-CoV-2 infection; SD, standard deviation. P-values are reported for comparison between males and females. Comparisons were performed using t-test, non-parametric rank sum test, or chi-square test as appropriate.
